# Associations Between Salivary HHV-6 And Long COVID Severity in a Cross-Sectional, Case-Control Analysis

**DOI:** 10.64898/2026.05.19.26353495

**Authors:** Claire S. Laxton, Alexandra Tabachnikova, Lily Cooke, Kexin Wang, Simone Blaser, Julio Silva, Jamie Wood, Henna S. Nam, Zhenni Lu, Christine Miller, Gisele Rodrigues, Victoria Fisher, Christian Guirgis, William B. Hooper, Alexandra Lee, Mackenzie Doerstling, Bornali Bhattacharjee, Leying Guan, David Putrino, Akiko Iwasaki

**Affiliations:** Department of Immunobiology, Yale School of Medicine, New Haven, CT; Department of Rehabilitation and Human Performance, Icahn School of Medicine at Mount Sinai, New York; Department of Biostatistics, Yale School of Public Health, New Haven, CT; Department of Medicine, Division of Infectious Diseases and Global Health, Yale School of Medicine, New Haven, CT; Department of Pediatrics, Division of Infectious Diseases and Global Health, Yale School of Medicine New Haven, CT; Center for Infection and Immunity, Yale School of Medicine, New Haven, CT; Howard Hughes Medical Institute, Chevy Chase, Maryland

**Author notes:** Co-first authors (Claire S. Laxton, Alexandra Tabachnikova, Lily Cooke, Kexin Wang).

**Keywords:** Long COVID, saliva, herpesviruses, hormones, symptom profiling, qPCR

## Abstract

**Background:** Reactivation of human herpesviruses (HHVs), particularly EBV, is associated with more severe acute SARS-CoV-2 infection and the development of Long COVID (LC). Reactivation of HHVs has been previously implicated in hormonal dysregulation, which has also been observed in LC, suggesting a potential link. This study aims to examine the relationship between LC symptoms and salivary HHV DNA, cortisol, testosterone, and oestradiol levels.

**Methods:** Our cross-sectional, case-control study enrolled 45 participants with LC, and 45 age-sex-matched controls. Surveys and validated health questionnaires were used to collect demographics, medical history, and symptom profiles. Saliva was self-collected at waking, 15, 30, and 45 minutes, and 8 and 16 hours after waking, across two consecutive days. Salivary cortisol, testosterone and oestradiol were measured, and extracted nucleic acid was tested for EBV, HSV 1/2, HCMV and HHV-6 A/B using multiplex qPCR, and for SARS-CoV-2 and RNaseP using RT-qPCR.

**Findings:** Detection of salivary EBV and HHV-6 DNA copies was highest early in the morning. There were no significant differences in salivary cortisol, testosterone, or oestradiol, or in EBV or HHV-6 DNA between the LC and control groups. However, salivary HHV-6 DNA levels were positively associated with a greater aggregated LC propensity score (r= 0.24, 95% CI= 0.02 to 0.44, p= 0.015, adj. p= 0.030), as well as anxiety (β= 0.69, 95% CI= 0.02 to 1.36, p= 0.045) and depression (β= 0.24, 95% CI= 0.05 to 0.42, p= 0.013) scores.

**Interpretation:** The observed correlation between salivary HHV-6 DNA and symptom severity suggests HHV-6 may contribute to post-acute disease, though mechanisms remain unclear. While we did not identify a relationship between salivary EBV DNA and LC, EBV may still play a role at earlier time points in the disease course, or in compartments not sampled in this study. These findings highlight the potential importance of HHV-6 in LC pathophysiology and underscore the need for longitudinal, multi-compartment studies of herpesvirus reactivation in LC.

**Funding:** This work was supported in part by the Else Kröner Fresenius Prize for Medical Research 2023 (to A.I.), the Howard Hughes Medical Institute Collaborative COVID-19 Initiative (to A.I.), and the Howard Hughes Medical Institute (to A.I.), the Polybio Research Foundation (to A.I. and D.P.) and the Steven and Alexandra Cohen Foundation (to D.P.). National Institutes for Health (NIH) funding sources supported AT (NIAID F31AI181508), SB (NHLBI T32HL007974-24) and CM (NCATS TL1 TR001864).

**Research in Context:** *Evidence before this study:* An estimated 400 million people have developed Long COVID, a debilitating disease in which symptoms persist after SARS-CoV-2 infection, potentially for years, affecting multiple organ systems. We searched PubMed for articles published from Jan 1, 2020, to Dec 31, 2025, using terms for long COVID (e.g., "long COVID", "PASC") combined with herpesviruses (e.g., "Epstein-Barr virus", "HHV-6") or hormones (e.g., "cortisol", "testosterone", "estradiol"), excluding reviews, editorials and comments, with additional relevant studies identified separately. From this, we identified multiple studies which found that reactivation of human herpesviruses (HHVs) during or shortly after acute SARS-CoV-2 infection is associated with an increased risk of developing Long COVID, although there were also studies showing no evidence of elevated HHV reactivation in Long COVID. Some studies found cortisol was lower in people with Long COVID, though others showed no differences. Separately, HHVs, particularly HHV-6 and EBV, have been implicated in hormonal dysregulation, suggesting there may be link between the two findings. However, most data were generated from single timepoint serum measurements of hormones or antibodies.

*Added value of this study:* Our study focused on individuals living with Long COVID for, on average, 2 years. We used saliva, which is easy to self-collect at multiple timepoints, to examine potential connections between HHV reactivation, hormonal disturbances, and Long COVID symptom severity. We collected and tested samples over two days for both HHVs and hormones and administered detailed health and symptom surveys. We did not observe differences in hormone levels between people with Long COVID and controls, although there were some trends between salivary HHV levels and hormones. We also did not observe clear differences in salivary EBV, CMV, HHV-6 or HSV DNA levels between controls and people living with Long COVID. However, we observed that elevated salivary HHV-6 DNA was associated with worse Long COVID symptom severity and potentially higher anxiety and depression scores, though the latter findings require further confirmation.

*Implications of all the available evidence:* Taken together with previous studies, our findings suggest that while EBV may play a role in the pathogenesis of Long COVID, before, during or shortly after acute infection, HHV-6 may have an ongoing pathological role. However, it remains unclear whether HHV-6 causes or exacerbates Long COVID symptoms or is merely an indirect indicator. Further studies are needed to paint a conclusive picture regarding the relationships between hormone disruption, HHV reactivation, and Long COVID.

## Introduction

There are nine human herpesviruses (HHVs). Most individuals become infected with one or more HHV during early life, typically with a mild primary infection ^1,2^. All HHVs, except varicella zoster virus (VZV), are shed in saliva during both primary infection and reactivation, which can occur without accompanying symptoms ^1^. Recently, HHV latency and reactivation have been associated with, or causally linked to, numerous chronic autoimmune disorders, including Multiple Sclerosis (MS) ^3–7^, Systemic Lupus Erythematosus (SLE) ^8,9^, Myalgic Encephalitis/Chronic Fatigue Syndrome (ME/CFS) ^10,11^, and other neurodegenerative diseases ^12–14^.

Almost as soon as SARS-CoV-2 emerged, HHVs, particularly EBV, have been implicated in both increased acute COVID-19 disease severity ^15–19^ and the development of the post-acute SARS-CoV-2 syndrome known as Long COVID (LC) ^20–26^. Su *et al.* demonstrated that EBV viremia during acute SARS-CoV-2 infection was associated with fatigue, memory problems, and sputum production in those who subsequently developed LC ^21^. Studies have also demonstrated that people with LC have elevated anti-EBV antibodies, indicative of a recent reactivation ^20,23,26^. Rohrhofer *et al.* detected EBV DNA in the throat washes of people with LC an average of 235 days after acute infection, at significantly higher rates than in recovered controls (50% vs 20%), although they did not observe differences in EBV-specific antibodies ^24^. Additionally, Zubchenko *et al.* demonstrated that individuals with LC who tested positive for reactivation of EBV and/or HHV-6 by PCR were more likely to experience sleep disorders, joint pain, hair loss and pulmonary symptoms ^22^. The authors did not distinguish the effects of EBV and HHV-6, nor HHV DNA detection in blood vs saliva, and thus, the precise relationships between observed elevations in oral HHV DNA levels and LC symptoms remain unclear.

Previous studies have also identified altered hormonal signalling in LC, including decreased blood cortisol levels ^23^ and lower salivary cortisol post-COVID, with or without Long COVID symptoms ^27^, particularly, as identified in a recent pre-print, during early morning timepoints ^28^. Moreover, LC is more prevalent in women, and sex-segregated analysis has highlighted different immunological profiles between males and females, and depending on menstrual cycle stage ^29–32^. However, prior studies looking at sex hormone levels in LC have used serum samples from a single timepoint.

HHV reactivation itself may also influence hormone secretion in Long COVID. Previous work showed that hair cortisol levels were reduced following acute EBV infection ^33^, and evidence from mouse studies has mechanistically linked HHV-6 with hypothalamic-pituitary-adrenal (HPA) axis disruption ^34^.

Saliva is non-invasive and simple to self-collect over many timepoints ^35^, and thus provides an opportunity to robustly interrogate diurnal variations in both stress and sex hormones ^36–43^, and HHV reactivation, as most HHVs are shed in saliva upon reactivation ^1^. Saliva has also previously been used to examine the relationships between HHVs and/or hormones and ME/CFS, a condition that shares similarities with Long COVID ^33,44^. Thus, this study aimed to intensively characterize the diurnal salivary dynamics of EBV, HSV, HCMV and HHV-6 DNA levels, and explore their associations with hormones and LC symptoms, to better establish the incidence and impact of chronic herpesvirus reactivation in people with LC.

## Methods

### Ethics

This study adhered to Declaration of Helsinki principles and was approved by the Mount Sinai Program for the Protection of Human Subjects (IRB 20-01758) and the Yale Institutional Review Board (IRB 2000029451). Written informed consent was obtained from all enrolled participants.

### Study design

This was a satellite study of the MY-LC (Mount Sinai - Yale Long COVID) project, a cross-sectional multi-site retrospective case-control study examining LC ^23^. This sub-study, which is reported in accordance with the STROBE guidelines for observational studies, was a repeated-measures, cross-sectional, case-control study in which participants were sampled over two consecutive days during the period of April 2023 to September 2024, with no further study visits. Participants were recruited by convenience sampling from the New York City area via social media posts and flyers posted throughout the Mount Sinai Health System. Participants with LC were also recruited from the Cohen Center for Recovery from Complex Chronic Illness and Center for Post COVID-19 Care at the Icahn School of Medicine at Mount Sinai, including individuals that have previously attended the clinic/centre. If the physician reviewing the patient identified that they met the inclusion/exclusion criteria, with the patient’s permission, they connected patients with the study coordinator for enrolment. Control participants were also recruited via emails to an existing database of individuals who received clinical home monitoring from the Abilities Research Center team during a past acute COVID-19 infection.

LC cases were included if they were aged ≥18 years old and were diagnosed with LC, which was initially defined as having symptoms persistent >6 weeks following initial COVID-19 infection, as the study was designed and enrolment began prior to the publication of consensus definitions of LC. Convalescent controls were included if they were aged ≥18 years old and reported either no previous COVID-19 infection or a complete recovery from COVID-19 within 6 weeks. Participants were excluded if they could not provide informed consent and had conditions preventing saliva collection. Post-enrolment, based on the subsequently published 2024 NASEM LC definition ^45^, the LC case definition was narrowed to having symptoms persisting for >3 months after initial SARS-CoV-2 infection. Therefore, cases were excluded if their initial SARS-CoV-2 infection was <3 months before enrolment and controls were excluded if they reported any SARS-CoV-2 infection <3 months before enrolment. Participants were also excluded if they reported a current or previous diagnosis of an autoimmune disease or immune deficiency, usage of any immunosuppressive or anti-herpes antiviral medication in the last 3 month, or if they did not meet age-sex matching criteria (matched reported sex and age, ±5 years).

### Participant surveys

One day prior to sample collection, demographic information and self-reported medical histories were collected using online surveys and further reviewed through examination of the electronic medical records (EMR) of collaborating clinics, or individual follow up, where required. Participants provided their gender on surveys, and biological sex was confirmed by EMR review or individual follow up. In all cases, as reported gender and sex matched, sex is reported hereon.

At the time of the initial medical and demographic survey, participants also completed the following validated health questionnaires: EuroQol 5-Dimension 5-Level health related quality of life (EQ-5D-5L) ^46^, Medical Research Council (MRC) dyspnoea scale (MRC dyspnoea) ^47^, Fatigue Severity Scale (FSS-7) ^48,49^, Patient Health Questionnaire (PHQ-2) ^50^, Generalized Anxiety Disorder 7 (GAD-7) ^51^, Self-report Leeds Assessment of Neuropathic Symptoms and Signs pain scale (S-LANSS) ^52^, Epworth sleepiness scale (ESS) ^53^, Single-Item Sleep Quality Scale (SQS) ^54^, Adult PROMIS Ability to Participate in Social Roles and Activities short form 8a (PROMIS Participation) ^55,56^, Neuro-QoL Adult Cognitive Function short form (NQ-Cognitive) ^57,58^, DePaul Symptom Questionnaire - Post-Exertional Malaise short form (DSQ-PEM) ^59^, and General Symptom Questionnaire-30 (GSQ-30) ^60^. Additionally, participants scored their overall health on that day (eq5), and global fatigue and breathlessness over the past week using 0–100-point visual analogue scales (VAS).

At the time of each saliva sample collection, participants were also asked to fill in a short online survey, detailed in **Supplementary Figure 1**. Here, participants were asked to rate their fatigue on a 0-100 VAS and provide a free-text answer to the question: “Excluding fatigue, what is the worst symptom you are experiencing at this exact moment?”. Answers were encoded into the end-organ systems using pre-specified criteria that had been agreed upon by two physicians (SB and CM, **Supplementary Document 1**). Encoding was completed twice for each survey, by two independent encoders (HN and ZL). Final encoding was independently reviewed by a physician (SB), who settled discrepancies and ambiguous answers, and any answer that was too ambiguous to be categorized was encoded as "Other." Additional encoding of neurocognitive symptom sub-types was completed in the same fashion. Sub-types were categorized as either fatigue (including references to tiredness, sleepiness or grogginess), head pain/pressure (included references to headaches, migraines, pressure in head), dizziness/light-headedness (including references to orthostatic symptoms), and brain fog (included all references to “brain fog”, issues with reasoning, memory, attention, and speed of thought). The term brain fog was selected as the umbrella term as it was the most frequent phrasing chosen by participants and is consistent with usage in previous studies ^61^.

Survey data was collected using REDCap and stored on Icahn School of Medicine, Mount Sinai servers. Once all data was collected, a copy of the de-identified database was transferred to researchers at Yale for analysis. Raw data from validated health questionnaires were aggregated and normalized according to their respective published protocols, as cited above.

### Sample collection and processing

Participants provided 12 saliva samples total over two days, with 6 samples per day self-collected at home by passive drooling into a 5 mL sterile Eppendorf tube. The collection schedule and restrictions are shown in **Supplementary Figure 1**. Samples were stored in home refrigerators then shipped by courier using cold blocks within 120 hours to Icahn School of Medicine, where they were stored at –80°C, before being shipped to Yale School of Medicine on dry ice and stored at – 80°C.

At Yale, de-identified saliva specimens were inventoried and managed using the unique alphanumeric codes assigned at the time of collection. Saliva samples from each participant were thawed simultaneously on ice, then centrifuged at 2500 xg for 15 minutes at 4°C. Supernatants were aspirated and aliquoted under enhanced BSL-2 conditions, then stored alongside remaining pellets at -80°C. For all downstream testing, as far as possible, matched pairs were run in the same batches.

### Salivary hormone quantification

Aliquots of saliva supernatant were shipped to Salimetrics, LLC (Carlsbad, CA, USA) on dry ice and analytes were measured for cortisol (5100), testosterone (5138), and oestradiol (5160) by ELISA in duplicate and then averaged.

For cortisol, assay sensitivity was 0.007 μg/dL, assay range was 0.012 – 3.0 μg/dL and Intra-assay coefficients of variation (CVs) were 4.6 – 6% ^62^. For testosterone, assay sensitivity was 1.0 pg/mL, assay range was 6.1 – 600 pg/mL and Intra-assay CVs were 4.6 – 9.85% ^63^. For oestradiol, assay sensitivity was 0.1 pg/mL, assay range was 1 - 32 pg/mL and Intra-assay CVs were 7.13 –7.45% ^64^. Analysis of saliva for cortisol and testosterone was conducted in three batches. Oestradiol analysis was conducted in a single batch.

To integrate analysis across batches, repeated samples from multiple timepoints for 7 different study participants across groups (LC and Control) were measured between Batch 1 and 2 (n=57 total) and for 5 different study participants across groups were measured between Batch 2 and 3 (n=53 total). A linear model was fit between Batch 1 and 2 and between Batch 2 and 3. No adjustments were made for demographic covariates because study participants were age- and sex-matched. A multiplicative coefficient (the slope of the linear regression line) was used to correct data, with Batch 2 as the reference. For testosterone, the intercepts against batch 2 were 3.93 and 10.47 for batch 1 and 3, respectively, and these were added to each value during correction. For cortisol, the intercepts were <0.01, so no additional additive correction was used.

Diurnal cortisol and testosterone analyses were conducted on natural-log-transformed values to reduce the right-skewness of the data distribution and mitigate the impact of outliers. For the Area Under the Curve (AUC) and Cortisol Awakening Response (CAR) AUC, individual natural log-transformed values for two consecutive days of each time point were averaged and then AUC was calculated. For participants without 2 samples per timepoint, the single timepoint was used in place of the average. AUC measures were calculated using the trapezoid method, with respect to ground and the waking timepoint for Whole Day AUC and CAR AUC, respectively, as described previously ^65^. For Waking testosterone, values were averaged between the first timepoint of two consecutive days. For oestradiol, a single morning timepoint value was measured.

### Salivary herpesvirus DNA quantification

As far as possible, 6 samples for each participant were tested: timepoints 0 (immediately after waking), 8 and 16 hrs after waking, from both days. Where a timepoint 0 sample was unavailable, for example due to insufficient sample volume, the next earliest morning sample available (e.g. 15-45 min after waking) was used instead.

### Saliva DNA extraction

Aliquots of 200 µL salivary supernatant were thawed in batches on ice and briefly spun. As far as possible, total nucleic acid (TNA) was extracted from 175 µL of each sample using the MagMax Viral Pathogens II kit (ThermoFisher Scientific, USA, catalogue: A48310) and KingFisher Apex (ThermoFisher Scientific, USA) as described previously ^66^, except using the MVP-II protocol, and including two elution steps, each into 50 µL elution buffer. Each 96-well extraction plate also included 6 negative extraction controls (nuclease-free water) interspersed throughout. Following elution, samples were immediately stored at -80°C.

### RT-qPCR and qPCR

The two assays, a multiplex qPCR to detect gDNA from the herpesviruses EBV, HSV 1 and 2, CMV and HHV-6 A and B, and a dualplex Reverse-Transcriptase (RT) qPCR to detect RNA from SARS-CoV-2 and TNA from human RNAseP, were run simultaneously on each batch of samples, as far as possible. Primers and probes for each assay were obtained from Integrated DNA Technologies (IDT, USA), and are listed, along with the final concentrations used, in **Supplementary Table 1**.

Both assays used 5 µL template per reaction, run in duplicate, in a final volume of 20 µL. Duplicate no-template control (NTC) wells using nuclease-free water were included on every plate. The herpesvirus multiplex used qPCR, Luna® Universal Probe qPCR Master Mix (Catalogue: M3004E, New England Biolabs, USA), with the following conditions: 95°C for 2 mins, [95°C for 15 secs, 60°C for 30 secs] * 40 cycles. For standard curves, the following quantitative herpesvirus gDNA standards were mixed 1:1:1:1 to make ten-fold dilutions ranging from 50,000-5 copies/µL: VR-3247SD, VR-538DQ, VR-1493DQ, VR-1467DQ (American Type Culture Collection, USA).

Standard curves analyses were done using simple linear regression with GraphPad Prism (Version 10.6.1). Efficiency was established using standard curves across a dynamic range of 10^1^-10^6^ copies/reaction conducted in triplicate (**Supplementary Figure 2A-B**). Lower limit of quantification (LLOQ) for each target was established with 12 replicates for each dilution over a range of 3-2187 copies/reaction (**Supplementary Figure 2C**). LLOQ was the lowest value which could be included in analysis of curves which meet the following criteria: amplification of 11/12 replicates with a Cq SD <1, efficiency (10^(-1/slope)-1^) of 95%-105%, R^2^ > 0.9800, and passing the replicates test for lack of fit (p>0.05). An individual was considered positive if a given virus was detected in ≥2 of their samples above the LLOQ.

For the SARS-CoV-2/RNASeP RT-qPCR, the Luna® Probe One-Step RT-qPCR Kit (No ROX) was used (catalogue: E3007E, New England Biolabs, USA), with the following conditions: 55°C for 10 mins, 95°C for 2min, [95°C for 30 secs, 55°C for 30 secs] * 45 cycles. The following positive controls were used at a single concentration: Hs_RPP30 positive control (IDT, USA) at 5,000 copies/µL and the Assay Ready Synthetic SARS-CoV-2 RNA Control (Catalogue: 103925, Twist Biosciences, USA) at 100 copies/µL. An individual was considered positive if SARS-CoV-2 amplified with a Cq <40 in ≥1 of their samples.

Thermocycling was done in two side-by-side CFX96 Touch (Bio-Rad, USA) machines, within the same calibration cycle. Following fluorescence baseline correction and manual inspection of the curves, data were exported as .csv files.

### Automated qPCR analysis pipeline

A custom Python3 extract-transform-load pipeline was created in-house for qPCR data analysis. This pipeline allows for rapid compilation and QC of multiple raw qPCR data files into one clean data sheet, ready for downstream analysis. The pipeline, which is now available for public use, and supporting files, are deposited in GitHub (https://github.com/clairelaxton/Automated_qPCR_ETL_pipeline).

Within the pipeline, data were first extracted from the raw .csv files exported from the Bio-Rad CFX96 qPCR machines, and sample IDs and targets automatically assigned according to user-supplied plate maps and fluorescent channel target assignments. Runs were then QC’d to flag negative controls (NTC and NEC) with amplification or positive controls without amplification, and that all samples had amplification for an internal control, here, RNAseP.

Next, the mean and SD for the Cycle of quantitation (Cq) and the Starting quantity (SQ), which were interpolated from the standard curve using the Bio-Rad CFX96 software prior to export, were calculated for each duplicate. Samples were then flagged as “Inconclusive” where they: contained replicates which amplified with Cq <10, had only one replicate which amplified and with a Cq >35, or had more than 1 replicate with a Cq SD ≤2. Finally, the mean SQ for each herpesvirus was normalized according to total input nucleic acid by multiplying by a correction factor, which was calculated for each sample by dividing the mean sample RNaseP Cq by the global median RNAseP Cq (adjustment for RNAseP or another internal control is a toggleable feature of the pipeline).

Where the pipeline marked samples ‘Inconclusive’, these were re-run a maximum of twice, and those that remained Inconclusive were excluded from analysis. Viral copies were finally converted to total copies/input in the reaction by multiplying by 5.

### Statistics

Overall data missingness due to insufficient sample or technical issues are reported in **Supplementary Figure 3A**. Statistical significance of differences in health questionnaire scores, hormone levels and viral copies between LC and control groups was assessed using Wilcoxon rank-sum test or signed-rank test, as indicated, as well as by regression modelling, described below.

Effect sizes for Wilcoxon tests were calculated as r, with 95% confidence intervals obtained via percentile bootstrap (2,000 resamples), using the R package rstatix ^67^.

Multiple testing correction was applied within focused subsets of related tests addressing the same biological question. Specifically, for panels including multiple viral outcomes, health questionnaire outcomes, or salivary hormones, p-values were adjusted within each relevant family of tests using Holm’s procedure to control the family-wise error rate (FWER) at 5% ^68^. Only tests of the primary hypotheses were included; covariates used to control for potential confounding were excluded. For stratified analyses, correction was applied separately within each participant group.

### Hierarchical symptom clustering

To assess whether participants cluster by symptom burden measured by the GSQ-30, we performed hierarchical clustering on the symptom severity profiles. Participant–participant dissimilarity was computed using Gower distance, treating symptoms as ordinal severity measures, while symptom– symptom distance was calculated from Spearman correlation. We then applied hierarchical clustering with average linkage to both the participant and symptom dimensions. The number of participant clusters was selected using the gap statistic: we chose k=3 because the gap increased sharply from k=2 to k=3, with only marginal gains for k>3 (**Supplementary Figure 3B**).

### Long COVID propensity score composition

We constructed a LC propensity score (LCPS) using a LASSO logistic regression model from the following 14 survey-based measures: EQ-5D-5L USA-indexed score, FSS-7 mean, PHQ-2 sum, GAD-7 sum, ESS sum, PROMIS Participation T-score, NQ-Cognitive T-score, DSQ-PEM sum, MRC dyspnoea, SQS, GSQ-30 sum, eq5 VAS, Fatigue VAS, Breathlessness VAS. This score followed the same conceptual framework as reported previously ^23^. All survey variables were standardized with LC status as the binary outcome. Predictive performance was evaluated using nested cross-validation, with 10 outer folds for performance estimation and 10 inner folds for model tuning, summarized by AUROC. We then refit the model on the full dataset and selected the final penalty parameter using the one-standard-error rule to favour a parsimonious solution. The surveys with non-zero coefficients in the final model are shown in **Supplementary Figure 3C**. For each participant, we then derived the model-based linear predictor as the LCPS, where higher values indicate survey profiles more strongly aligned with LC. We assessed the association between LCPS and HHV-6/EBV and salivary hormone levels using Spearman correlation and linear regression models adjusted for age, sex, and BMI with Holm-adjusted p-values. Spearman correlations between herpesvirus copies and hormones were also performed. Confidence intervals for spearman correlations were calculated with Fieller correction using the R package correlation ^69^.

### Modelling EBV and HHV-6 salivary DNA detection across time

We analysed longitudinal viral measurements using a Bayesian mixed-effects model. The 2-day average for each timepoint and participant was taken. Values below the lower limit of quantification (LLOQ = 3) were treated as left-censored at LLOQ. We then fit a Bayesian lognormal mixed-effects model using R package brms ^70^ with fixed effects for LC group, age, sex, BMI, and categorical time points, along with a participant random intercept, and we reported posterior means and 95% credible intervals.

### Regression modelling of symptoms, viral copies and hormones

Replicate virus measurements were averaged within each participant at the same time point across the two study days. We used linear mixed-effects models (lmer in R package lme4 ^71^) to account for repeated measures per participant. We reported all regression coefficients with 95% confidence intervals and highlighted associations with a nominal p<0.05. Within each model, Holm correction was applied to all EBV and HHV-6 p-values.

Fatigue was modelled as the outcome, with fixed effects for categorical relative time to the first measurement, LC group, HHV-6, EBV, age, sex, and BMI, and a random intercept for participant ID. We then fit the same model separately within the LC and control subgroups to estimate within-group associations.

For each salivary hormone outcome, we fit linear regression models with mean log (HHV-6 or EBV +1) as predictors, adjusting for age, sex, BMI, hormonal medication use, and time since first SARS-CoV-2 infection (binned as <2 years or >2 years); in the combined cohort we additionally adjusted for LC versus control group. We also fit stratified models separately in LC and control groups as well as by sex.

For each validated health questionnaire outcome, we fit linear regression models with mean log (HHV-6 or EBV +1) as predictors, adjusting for age, sex, and BMI; in the combined cohort we additionally adjusted for LC versus control group. We also fit stratified models separately in LC and control groups. Due to their inclusion in the LCPS analysis, the three VAS questions (fatigue, breathlessness and general health-eq5) were also included in these analyses.

## Results

### Participants with Long COVID report an overall high disease burden across multiple metrics

In total, 160 participants were enrolled in the study (92 LC, 68 Controls) and following post-enrolment exclusions, as summarised in **Figure 1A**, 90 participants (45 age-sex matched pairs) were included for our analyses. Participants’ demographics, self-reported medical history, and medication usage are shown in **Figure 1B-E** and **Supplementary Table 2**. There were no statistically significant differences in BMI, days since last COVID vaccine dose, or days since last SARS-CoV-2 infection (**Figure 1B-E**). Prior COVID-19 incidence, levels of medication use, and co-morbidities were higher in the LC group.

**Figure 1:**
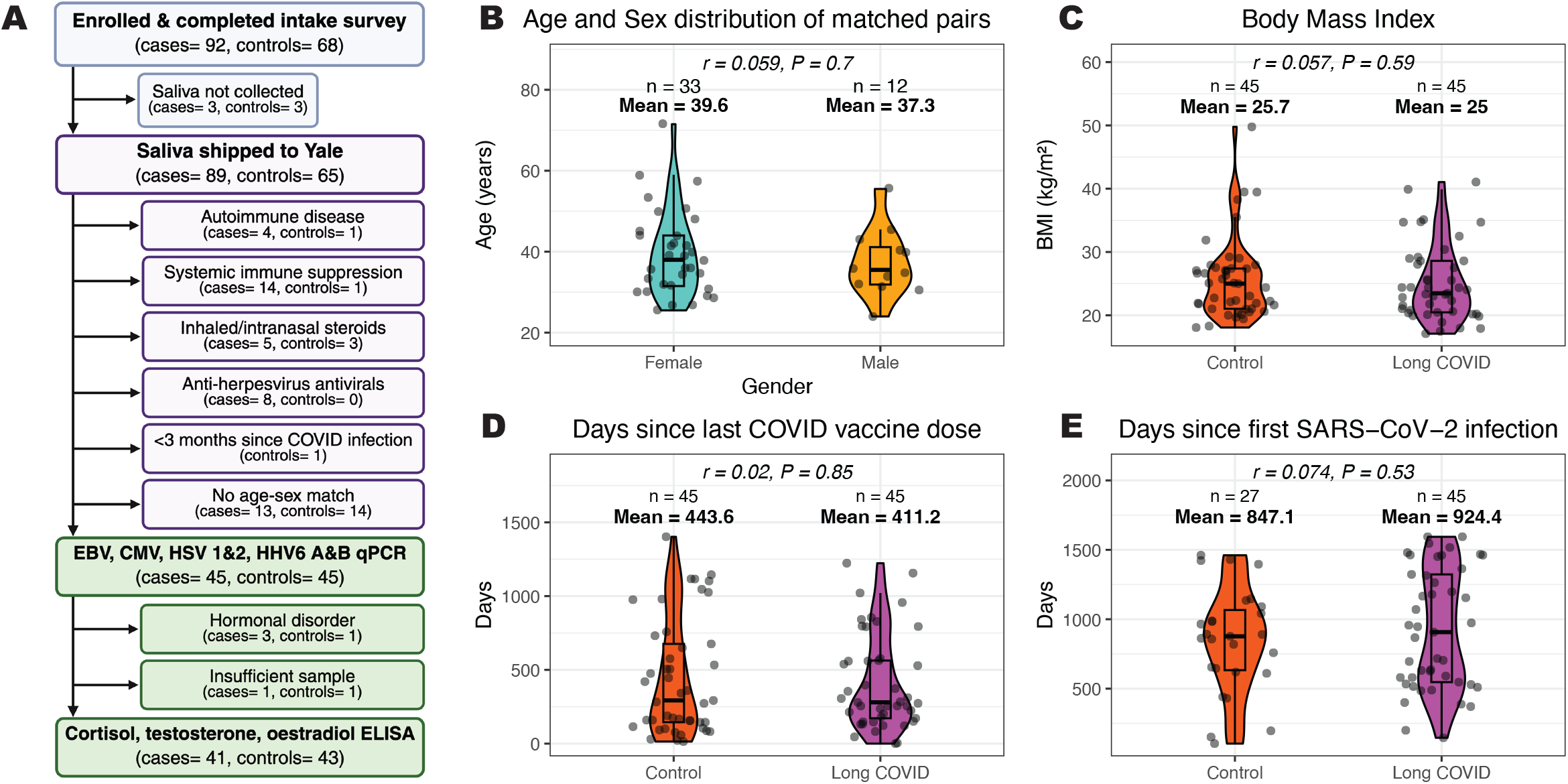
Cohort description and demographics. **A.** Flow chart of participant inclusion in the study. Cases defined as people self-reporting a Long COVID diagnosis, Controls are defined as those who reported never experiencing or being fully recovered from COVID-19. **B-E.** Description of MY-LC Saliva cohort characteristics. Differences between groups determined by Wilcoxon signed-rank test, p<0.05 was considered significant.

Between-group comparisons of validated health questionnaire outcomes, using paired Wilcoxon signed-rank tests, revealed significant differences. All outcomes had a Holm-adjusted p < 0.001 and are reported along with confidence intervals, in **Table 1**. Overall, compared with convalescent controls, those with LC reported severely reduced quality of life (EQ-5D-5L r= -0.87), cognitive function (NQ-Cognitive r= -0.85), and ability to participate in daily activities (PROMIS Participation r= -0.86) as well as more severe fatigue (FSS-7 r= 0.86), post-exertional malaise (DSQ-PEM r= 0.87), breathlessness (MRC dyspnoea r= 0.87) and general symptom burden (GSQ-30 r= 0.87). Additionally, those with LC reported moderately reduced sleep quality (SQS r= -0.57) and increased daytime sleepiness (ESS r= 0.58), as well as increased symptoms of anxiety (GAD-7 r= 0.71), and depression (PHQ-2, r= 0.75).

**Table 1:** Summary statistics from participant health questionnaires.

| Variable | N LC /Control | N pairs tested | LC (mean, SD) | Control (mean, SD) | adj. p-value | r | 95% CI |
| --- | --- | --- | --- | --- | --- | --- | --- |
| EQ-5D-5L, USA-indexed score | 45/45 | 45 | 0.6 ± 0.3 | 0.9 ± 0.1 | <0.001 | -0.87 | [-0.87, -0.85] |
| FSS-7 mean | 45/45 | 45 | 5.5 ± 1.6 | 1.7 ± 0.9 | <0.001 | 0.86 | [0.83, 0.87] |
| PHQ-2 sum | 45/45 | 45 | 2.1 ± 1.4 | 0.5 ± 0.9 | <0.001 | 0.75 | [0.58, 0.86] |
| GAD-7 sum | 45/45 | 45 | 7.6 ± 5.5 | 3 ± 3.1 | <0.001 | 0.71 | [0.53, 0.82] |
| S-LANSS sum | 27/0 | 0 | 8 ± 5.7 | NA | - | - |  |
| ESS sum | 45/45 | 45 | 7.2 ± 4.3 | 3.7 ± 2.9 | <0.001 | 0.58 | [0.36, 0.76] |
| PROMIS Participation T-score | 45/45 | 45 | 36.9 ± 7 | 59.3 ± 7.2 | <0.001 | -0.86 | [-0.87, -0.84] |
| NQ-Cognitive T-score | 45/45 | 45 | 38.8 ± 8 | 54.5 ± 7.4 | <0.001 | -0.85 | [-0.87, -0.81] |
| DSQ-PEM sum | 45/45 | 45 | 28.8 ± 20.5 | 0.3 ± 0.9 | <0.001 | 0.87 | [0.87, 0.87] |
| MRC dyspnoea | 45/45 | 45 | 2.4 ± 1 | 1.1 ± 0.3 | <0.001 | 0.87 | [0.82, 0.90] |
| SQS | 45/45 | 45 | 4.4 ± 2.1 | 6.8 ± 2.1 | <0.001 | -0.57 | [-0.74, -0.33] |
| GSQ-30 sum | 44/44 | 43 | 48.4 ± 21 | 6.4 ± 5.9 | <0.001 | 0.87 | [0.87, 0.87] |
| eq5 VAS | 45/45 | 45 | 49.9 ± 20.3 | 86.2 ± 8.4 | <0.001 | -0.87 | [-0.87, -0.87] |
| Fatigue VAS | 45/45 | 45 | 63.4 ± 18.6 | 14.6 ± 15.6 | <0.001 | 0.86 | [0.83, 0.87] |
| Breathlessness VAS | 45/45 | 45 | 35.8 ± 22.9 | 5.8 ± 15.7 | <0.001 | 0.8 | [0.69, 0.87] |

Differences were evaluated by Wilcoxon signed-rank tests. Holm-adjusted (adj.) $p < 0.05$ was considered significant. Effect size $r$ is signed such that positive values indicate higher scores in the LC group, with 95% confidence intervals obtained via percentile bootstrap (2,000 resamples). LC = Long COVID.

Hierarchical clustering analysis of the GSQ-30 data using Gower distance, shown in **Figure 2A**, revealed that symptom severity clustered into low (n=45 Controls, n=14 LC), medium (n=25 LC) and high (n=5 LC) burden groups. At the time of each sample collection, participants rated their fatigue and recorded the single worst symptom they were experiencing, as well as its severity using a 0-100 VAS. These symptoms were encoded into end-organ-system categories and further classified according to whether pain was a feature (**Figure 2B**). Neurocognitive symptoms were most frequently reported, primarily headache/pressure and brain fog in the Long COVID group (**Supplementary Figure 3D**), followed by musculoskeletal and pulmonary symptoms. Those with LC reported significantly higher levels of fatigue (**Figure 2C**). Overall, participants also reported their lowest fatigue during the day, 8 hours after waking (8 vs. 0 hours post waking: β = -7.97, 95% CI= -11.78 to -4.15, p<0.001). When stratified by LC status, in the control group only, higher fatigue was associated with being female (β= 17.3, 95% CI= 5.72 to 28.87, p= 0.004) and having a higher BMI (β= 0.91, 95% CI= 0.10 to 1.71, p= 0.029).

**Figure 2:**
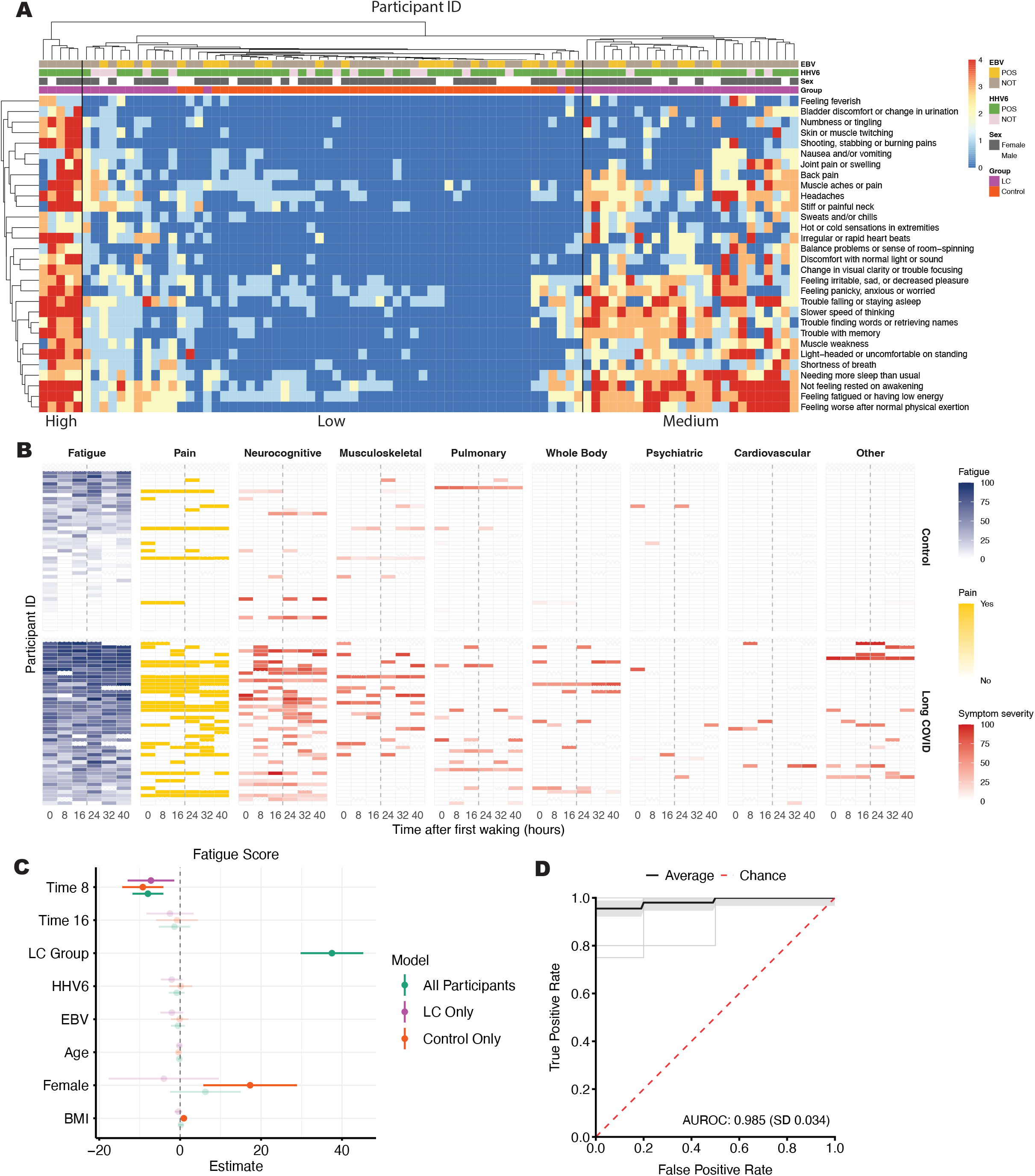
Analysis of symptom survey responses. **A.** Hierarchical clustering analysis of General Symptom Questionnaire (GSQ-30), using Gower distances (LC= Long COVID). EBV and HHV-6 positivity and sex are also indicated. n = 88 (44 LC, 44 control). **B.** Heat map showing self-reported fatigue, pain and the worst reported symptom, encoded by end-organ-system, at each sample collection timepoint. **C.** Linear mixed-effects models, modelling fatigue at a given time point (as in panel **B)**, with fixed effects for timepoint (categorical, 2-day average timepoint, using 0 hours is the reference), LC group, HHV-6, EBV, age, sex, and BMI, and a random intercept for participant ID. Models were fit within LC and Control subgroups. Regression coefficients are reported with 95% confidence intervals, and those in bold are associations with p<0.05. **D.** Receiver Operating Characteristics (ROC) curve to assess predictive performance of the LASSO model to generate the Long COVID Propensity Score (LCPS).

To produce a quantifiable metric of symptom magnitude and prevalence, survey responses were compiled to produce a single metric, the Long COVID Propensity Score (LCPS). This score was generated using a LASSO-penalized logistic regression model, following the same conceptual framework previously described for MY-LC study participants ^23^. The LCPS performed very well in differentiating the LC group from Controls (AUC = 0.985, SD = 0.034, **Figure 2D**). The overall LCPS disease severity for each symptom cluster defined in **Figure 2A** is shown in **Supplementary Figure 3E**.

### There were no differences in salivary cortisol, testosterone or oestradiol between Long COVID and control groups

Approximately 20% (n=20) of participants, all female, reported taking hormonal medication. Hormonal contraceptives, including hormonal intrauterine devices, were the most reported hormonal medication, and hormonal medication use was balanced across LC and Control groups (**Supplementary Table 2**). We performed sensitivity analysis, comparing differences in hormone levels either including or excluding individuals who reported taking hormonal medications. This analysis showed that hormonal medications did not significantly affect hormone levels (data not shown). Additionally, 4 individuals self-reported hormonal disorders: hyperthyroidism (n= 1), polycystic ovary syndrome (PCOS, n=2), and hypothyroidism plus PCOS (n=1). Therefore, for our hormone analyses, we excluded these 4 participants with hormonal disorders and adjusted our linear models for hormonal medication use.

To capture diurnal dynamics of hormone production, salivary cortisol and testosterone were measured at 4 morning time points (in 15-minute increments from waking), an afternoon time point (8 hours after waking), and an evening time point (16 hours after waking) over two consecutive days using ELISA. Salivary oestradiol was measured at a single morning time point to capture the morning peak, while also conserving sample volume.

Cortisol peaked 15-30 minutes after waking and was secreted at similar levels in participants with LC and controls (**Figure 3A**). To quantify total Cortisol Awakening Response (CAR), AUC was measured from waking levels to 45 minutes, and it did not differ between LC and Control (**Figure 3B**). Total cortisol production over the course of the day measured with AUC trended lower in LC than Control but did not reach significance (p= 0.076, r= 0.19, 95%CI= 0.02 to 0.41, **Figure 3C**).

**Figure 3:**
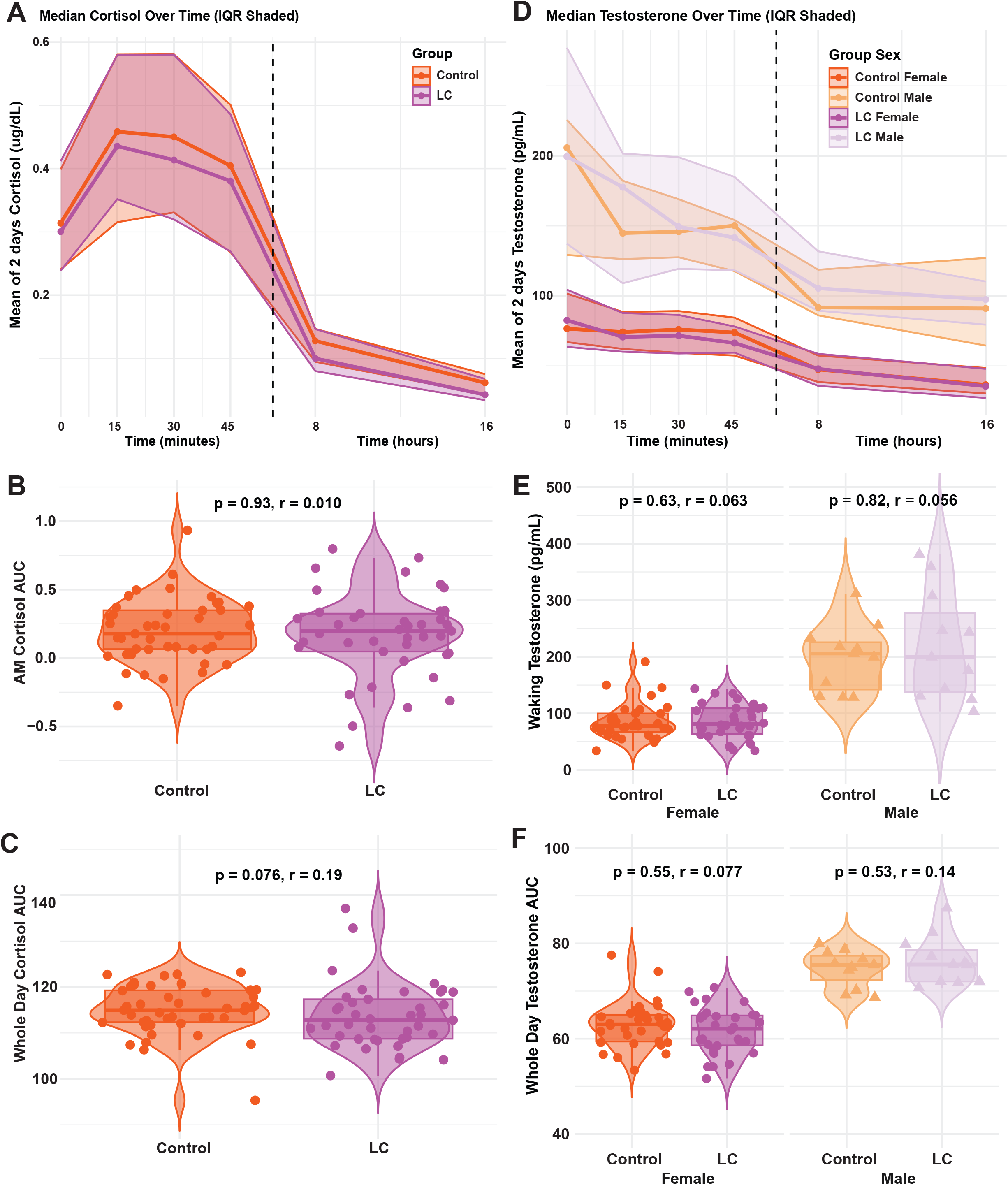
Salivary Hormones are not significantly different in Long COVID. **A, D.** Line plot showing median and Interquartile Range of 2-day mean of salivary Cortisol (**A**) and Cortisol (**D**) for each timepoint by group. Dashed vertical line indicates split of morning cortisol from afternoon and evening. **B, C.** Comparison of Area Under the Curve of Cortisol Awakening Responses for the first 4 timepoints, (**B**) and Area Under the Curve of Whole Day Cortisol for 6 timepoints, (**C**), averaged over 2 days by group. n=41 (LC) and n=43 (Control). **E, F.** Comparison of 2-day average of Waking Testosterone at the first timepoint (**E**) and Area Under the Curve of Whole Day Testosterone for 6 timepoints, averaged over 2 days, (**F**), by group. n=30 (LC Female), n=11 (LC Male), n=32 (Control Female), and n=11 (Control Male). For the violin boxplots, central lines indicate the group median values, and the top and bottom lines indicate the 75^th^ and 25^th^ percentiles, respectively. The whiskers represent 1.5 x the interquartile range. Each dot represents one individual. Statistical significance of the difference in median values was determined using Wilcoxon ranked-sum tests.

Testosterone, too, showed a diurnal production pattern, peaking upon waking and decreasing throughout the day, with higher levels in males (**Figure 3D**). However, testosterone was produced at similar levels in LC and Controls, both at waking (**Figure 3E**) and throughout the whole day as measured with AUC (**Figure 3F**). In both groups, males produced significantly more testosterone than females (p <0.001, **Figure 3E-F**). Similarly, morning oestradiol levels did not differ by group, but had a greater range of production in females, likely reflecting variations related to menstrual cycle (**Supplementary Figure 4F**).

When analysed using a linear model, hormone levels were not associated with LC status or herpes viral DNA, when adjusted for age, sex, BMI, time from infection and hormonal medication use (**Supplementary Figure 4A-E**). When stratified by sex, there were no significant associations between hormone level and LC status (**Supplementary Figure 4G-K**), though there was a small, non-significant trend toward lower whole-day testosterone in the LC group in the Female-only model (β= -2.20, 95% CI= -4.67 to 0.27, p= 0.080, **Supplementary Data 1**). Hormone levels also did not correlate with LCPS (**Supplementary Figure 4L-O**).

### Detection of neither EBV nor HHV-6 DNA was singularly predictive of Long COVID status

Saliva samples were tested for EBV, HSV 1 and 2, CMV, HHV-6 A and B DNA by qPCR, and SARS-CoV-2 RNA by RT-qPCR. An average of 6.0 samples per Control and 6.2 samples per participants with LC were tested, from 0, 8, and 16 hours after waking over two consecutive days.

EBV and HHV-6 positivity rates in Controls were 42.2% and 80.0%, respectively, and 24.4% and 77.8%, respectively in participants with LC (**Figure 4A**). Co-presence of EBV and HHV-6 was observed in 40% of Control participants and 20% of participants with LC. Additionally, no participants were positive for SARS-CoV-2 RNA, or HCMV or HSV-1/2 DNA.

**Figure 4:**
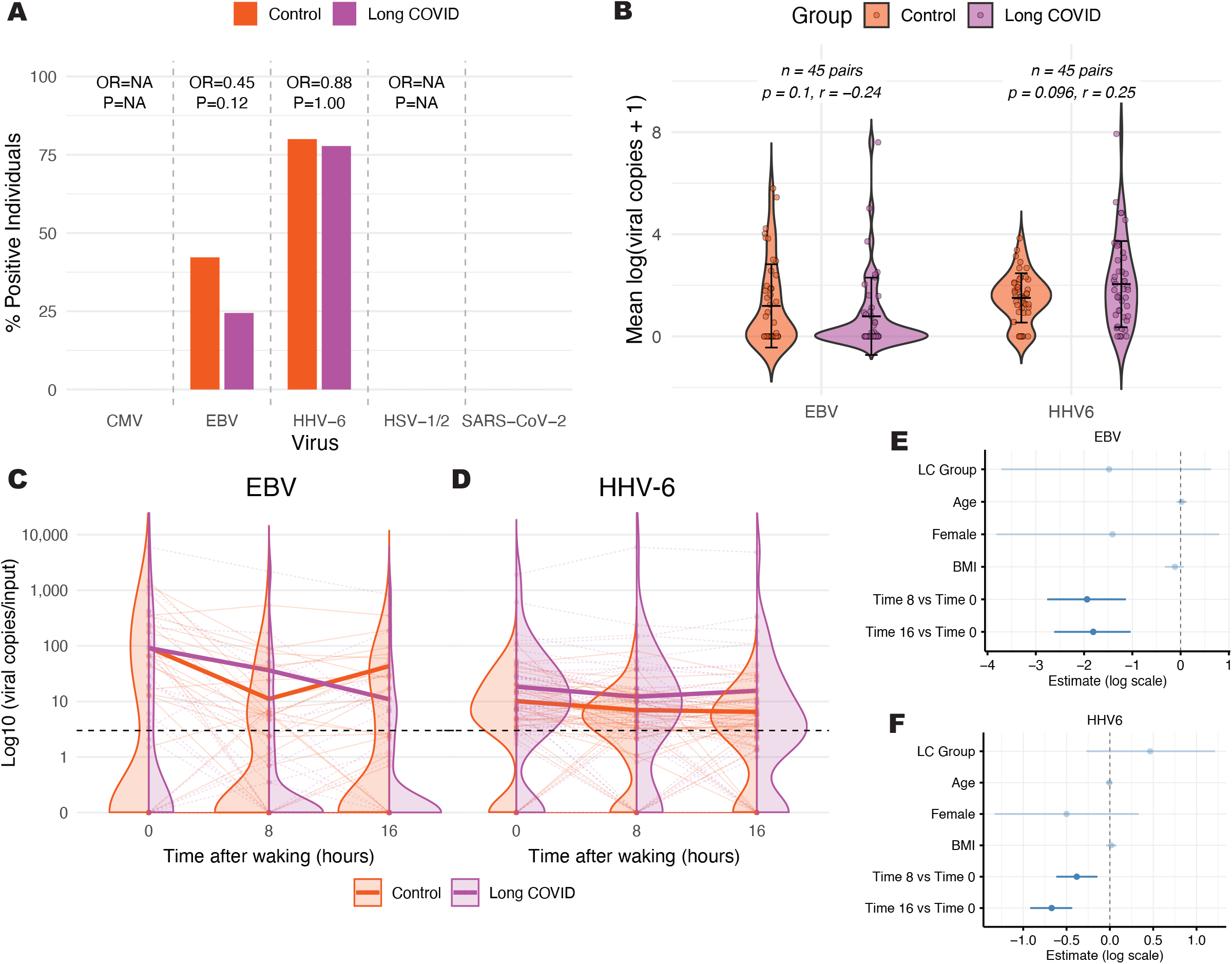
Detection of salivary herpesviruses. **A.** Positivity rate of all participants for each virus. Statistical differences tested using Fisher’s Exact test, OR= Odd’s Ratio, P<0.05 is significant. **B.** Comparison of mean log (viral copies+1) between participants with Long COVID and controls. Mean and SD are shown. Statistical differences tested using Wilcoxon signed-rank test, effect size based on the mean difference, p<0.05 is significant. **C-D.** Split violin plots showing viral copies of EBV and HHV-6 detected for each timepoint, averaged from 2 days. Bold line shows median of all samples >LLOQ (dashed black line). **E-F.** Forest plots from a Bayesian left-censored log-normal mixed model, showing associations between EBV and HHV-6 mean log (viral copies) +1, respectively, with timepoint and Long COVID status, adjusted for age, sex and BMI. Posterior mean and 95% credible intervals are reported, bold lines are those with credible intervals that do not cross zero.

There were no significant differences in viral positivity (**Figure 4A**) or proportion of positive samples per participants between groups (**Supplementary Figure 5A**). Similarly, although there was a trend toward higher mean HHV-6 DNA (r= 0.246, 95% CI= 0.02 to 0.51, p= 0.096) and lower mean EBV DNA (r= -0.241, 95% CI= -0.52 to -0.02, p= 0.099) copies in the LC group, the differences were not statistically significant (**Figure 4B**). There were no differences in area under the curve across all time points for either virus (**Supplementary Figure 5B**). Additionally, EBV and HHV-6 viral copies were not significantly correlated with each other (**Supplementary Figure 5C**).

Two-day salivary detection of EBV and HHV-6 DNA for each participant is shown in **Figure 4C-D**. Analysis using a Bayesian left-censored log-normal mixed model found that detection of higher levels of EBV and, to a lesser extent HHV-6, DNA was more likely at the first timepoint of the day compared to later timepoints (EBV 8 vs. 0 hours post waking: Estimate = -1.94, 95% CI= -2.76 to -1.13; EBV 16 vs 0 hours post waking: Estimate = -1.81, 97.25% CI= -2.62 to -1.04; HHV-6 8 vs. 0 hours post waking: Estimate = -0.38, 97.25% CI= -0.62 to -0.14; HHV-6 16 vs 0 hours post waking: Estimate = -0.67, 97.25% CI= -0.92 to -0.43; **Figure 4E-F, Supplementary Data 1**). This model also supported the non-significant associative trends in viral copy number and Long COVID status.

### Higher salivary HHV-6 DNA was associated with worse overall Long COVID symptom severity, anxiety, and depression

Next, we examined associations between salivary viral DNA and LC symptoms. When comparing the GSQ-30 clusters derived from **Figure 2A**, HHV-6 differed across groups (one-way ANOVA, p = 0.04). After adjusting for demographics, those with LC in the High and Medium severity groups had higher salivary HHV-6 DNA copy numbers than controls (**Figure 5A**, High vs. Control: β= 1.55, 95% CI= 0.27 to 2.84, p = 0.018; Medium vs. Control: β= 0.69, 95% CI= 0.01 to 1.36, p= 0.047) though only the effects in the High severity group survived multiple testing correction (**Supplementary Data 1**). Among those with LC, when analysed using a linear model with LC severity clusters as ordered levels, HHV-6 showed a positive monotonic trend across ordered LC clusters (Low → Medium → High), although the linear trend did not reach significance (p = 0.075), likely due to limited sample size within LC clusters.

**Figure 5:**
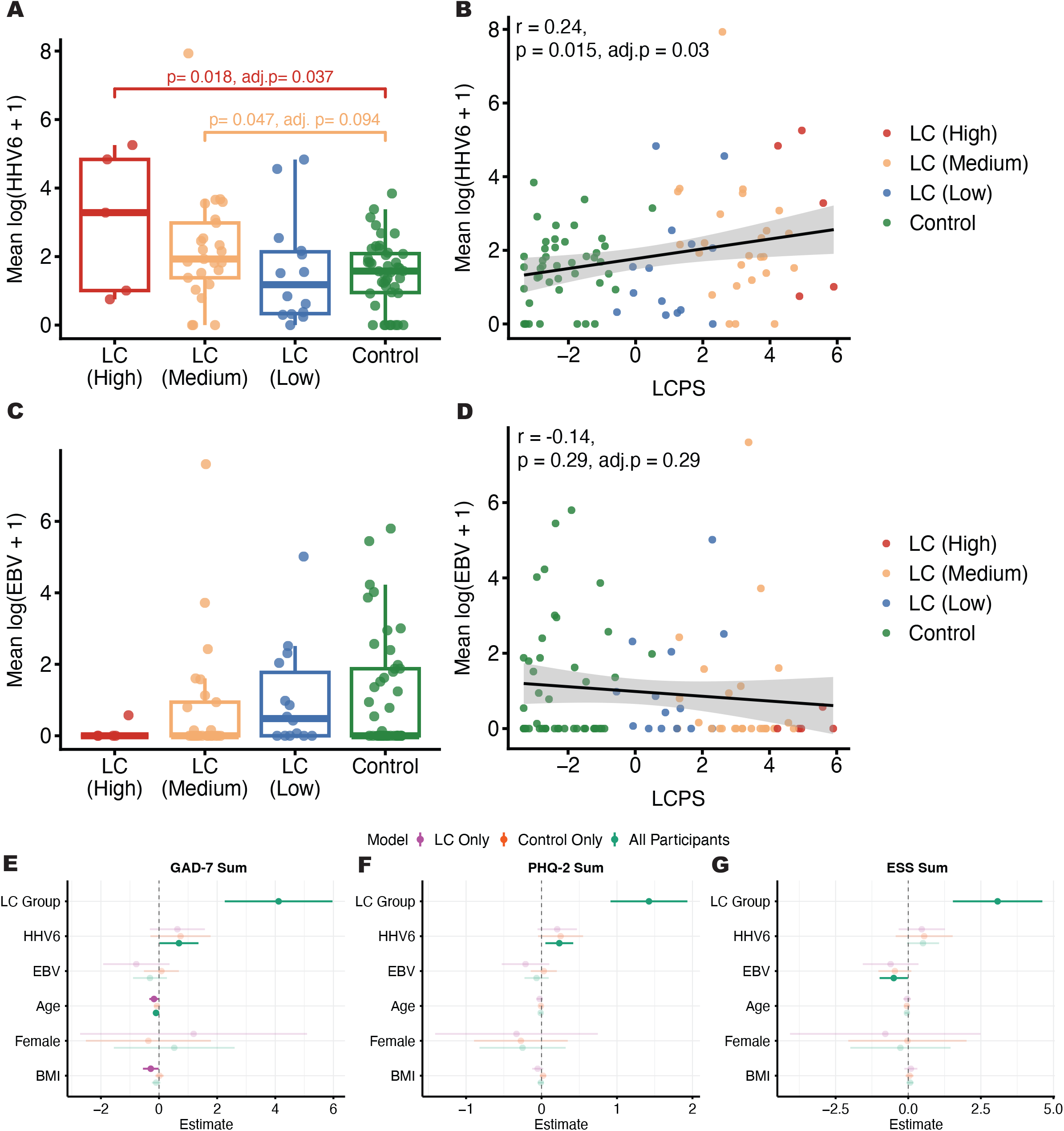
Relationships between salivary herpes DNA detection and Long COVID symptom severity. **A, C.** Boxplot showing HHV-6 (**A**) or EBV (**C**) DNA copies split according to GSQ-30 symptom cluster. Differences were tested using linear regression model with Control as the reference, adjusted for age, sex and BMI, nominal p<0.05 is significant, n = 88 (44 LC, 44 control). **B, D.** Correlation analysis between Long COVID propensity score and HHV-6 (**B**) and EBV (**D**) DNA copies, Spearman coefficients (R) are shown, significant determined by linear regression adjusting age, sex and BMI, Holm-adjusted (adj.) p <0.05 was considered significant, n = 88 (44 LC, 44 control). **E-G.** Forest plots showing the association between anxiety (GAD-7, **E**), depression (PHQ-2, **F**) and daytime sleepiness (ESS, **G**) scores and Long COVID status, EBV and HHV-6 mean log (viral copies) +1, adjusted for age, sex and BMI, and stratified by Long COVID status. Regression coefficients are reported with 95% confidence intervals and those in bold are associations with a nominal p<0.05. Nominal significance did not survive Holm’s multiple correction.

We then examined the relationship between salivary viral DNA copies and LCPS score. We found that among all participants, higher salivary HHV-6 DNA copy numbers were positively correlated with higher LCPS (i.e., worse severity) (**Figure 5B**, r= 0.24, 95% CI= 0.02 to 0.44, p= 0.015, adj. p= 0.030). However, there were neither significant differences in salivary EBV DNA copies between GSQ-30 symptom cluster group (**Figure 5C**), nor any significant correlations between LCPS and levels of salivary EBV DNA (**Figure 5D**).

To examine the relationships between salivary viral DNA detection and individual symptom types, we used a linear model to conduct exploratory analysis of the effect of salivary HHV-6 and EBV DNA copies on health questionnaire outcomes. Higher HHV-6 copies were associated with increased GAD-7 (anxiety, β= 0.69, 95% CI= 0.02 to 1.36, p= 0.045) and PHQ-2 (depression, β= 0.24, 95% CI= 0.05 to 0.42, p= 0.013) scores in all participants (**Figure 5E-F**), and higher EBV copies were associated with lower ESS (daytime sleepiness, β= -0.5, 95% CI= -0.99 to -0.01, p=0.045) in all participants (**Figure 5G**). However, these effects did not survive multiple testing correction, and further research is needed to be conclusive.

Finally, we examined the associations between salivary hormones and either EBV or HHV-6 DNA. Those with higher waking testosterone were more likely to have higher levels of EBV DNA in their saliva, which was predominantly driven by males in the LC group (**Supplementary Figure 6C,** waking testosterone, r= 0.32, 95% CI = 0.01 to 0.58, p= 0.041). This was supported by results from linear regression models which found a positive association between EBV and waking testosterone in all participants (β= 8.62, 95% CI= 1.75 to 15.49, p= 0.015) and in the LC-only model (β= 14.25, 95% CI 1.59 to 26.91, p= 0.029), (**Supplementary Figure 4B**) and in males only when stratified by sex (β= 26.92, 95% CI= 0.16 to 53.68, p= 0.049, **Supplementary Figure 4H**). However, these findings, both from the correlations and models, did not survive multiple testing correction.

## Discussion

This exploratory observational study aimed to characterize the relationship between herpesvirus reactivation, salivary hormones, and LC symptoms in patients ∼2 years after initial SARS-CoV-2 infection. We observed that those with LC reported significantly worse symptoms, across a wide range of metrics.

The levels of salivary hormones we observed align with those observed previously ^72–77^. However, there were no significant relationships between salivary cortisol, testosterone or oestradiol and LC status or LCPS. Although we observed a trend towards lower whole-day salivary cortisol in those with LC, this was not statistically significant. These observations contrast with previous findings by Klein et al. of lower serum cortisol in people with LC ^23^. The discrepancy may be explained by differences in measurement of free vs. bound hormone between serum (total) and saliva (free) ^38,40^, the viral variant that triggered LC, vaccination status, or by the fact that participants were enrolled ∼2 years after their initial infection in this study, a much later time from infection than Klein et al. A previous study described lower morning salivary cortisol in individuals post-SARS-CoV-2 infection, compared to previously uninfected individuals, but no differences between Long COVID and recovered individuals ^27^. Other studies have similarly found no differences in serum cortisol between Long COVID and control groups ^78^ . In our study, most control participants were previously infected with SARS-CoV-2; further comparison with validated ‘never infected’ controls is necessary to determine whether SARS-CoV-2 infection itself affects cortisol diurnal rhythm, and for how long any effect may persist.

We observed an overall salivary viral DNA positivity rate of 33% for EBV and 79% for HHV-6 across the study cohort, in line with observations from control groups in previous studies ^79,80^. However, other studies report lower rates of HHV-6 positivity in saliva ^81,82^, and EBV detection rates of up to 60% ^82^, suggesting a high level of variability in the literature, likely driven by differences in methodology, geography, and shedding kinetics. No individuals in our study were positive for SARS-CoV-2, HCMV or HSV-1/2 during sample collection.

Additionally, we observed that salivary detection of EBV, and to a lesser extent, HHV-6 DNA copies were significantly higher immediately after waking compared with later timepoints. While this has been observed previously for EBV ^83^, to our knowledge it has not been reported for HHV-6. This observation may be due to accumulation during sleep. However, we adjusted for host DNA/RNA quantity, suggesting that higher detection in the morning may be independent of sample ‘richness’ ^84^.

Previous studies have reported elevated antibodies against lytic antigens of EBV ^20,23^ in patients with LC, and EBV and/or HHV-6 positivity at the post-acute stage was associated with LC symptoms ^22^, suggesting that recent or ongoing reactivation may be driving disease. In this study, we observed no significant relationship between the levels of either HHV-6 or EBV DNA in saliva and LC status. This suggests that previous findings of elevated antibodies against EBV likely reflect reactivation of EBV either earlier in the disease process of LC ^23^ or during acute infection, as has been observed in the blood ^21^ and in the nasal mucosa ^19^. It is worth noting that several studies did not observe elevated EBV antibodies in Long COVID ^24,78,85^. Longitudinal investigation of viral production and circulating antibodies is required to determine this relationship more conclusively.

However, comparing a binary outcome-LC or Control - is somewhat crude given the heterogeneity of LC manifestations. In reality, participants’ symptomology fell on a spectrum, with a very small ‘high’ symptom burden group (n=5), and with overlap between those in the ‘low’ LC symptom burden group and Controls (**Figure 2A**). To account for this, we created the LCPS score as a continuous measure of LC severity. Here, we observed that having higher levels of salivary HHV-6, but not EBV, DNA was correlated with both a higher LCPS, and a greater symptom burden, being elevated in participants in the medium-to-high LC severity groups. Zubchenko et al. (2022) also observed a positive association between EBV and/or HHV-6 DNA and Long COVID symptoms, although they did not stratify their analysis by virus ^22^. Our study suggests that those findings may have been driven more by HHV-6 than EBV. However, we cannot rule out EBV reactivation occurring at a different body site, which we could not detect using saliva.

Previous work examining other neurological disorders aligns with our findings. For example, HHV-6 genomic sequences in blood samples were found to be more frequent in people with ME/CFS than healthy donors and higher copies were associated with a higher disease severity ^86^. Another study showed that detection of HHV-6 reactivation was greater in the blood of people with ME/CFS and MS ^87^. In a longitudinal analysis of HHVs in the saliva of people with ME/CFS, authors reported higher levels of salivary HHV-6B and HHV-7 DNA, which correlated with symptom severity in a subgroup of more severely symptomatic participants ^44^. Notably, other studies have found relationships between ME/CFS disease and EBV, not with HHV-6 ^88^. Both viruses likely play important but distinct roles in ME/CFS pathology. While we did not have sufficient data to determine the number of participants in our Long COVID group which may also meet the diagnostic criteria for ME/CFS, it is likely that many would, given the greater overall DSQ-PEM frequency and severity scores in the Long COVID group.

Further exploratory analysis found a positive association between salivary HHV-6 DNA and anxiety and depression. HHV-6 has wide tropism, including olfactory astrocytes, which are a major site of latency and a likely mode of entry into the central nervous system ^34,89–91^. Detection of HHV-6 in the Purkinje cells of human post-mortem cerebellar tissue was associated with major depressive disorder (MDD), bipolar disorder, and suicide ^90^. People with MDD also had higher antibody titres against the latent HHV-6B protein SITH-1 ^91^. Anti-SITH-1 antibodies were also elevated in people with LC, and seropositivity was associated with more severe fatigue and depression scores ^92^. In a mouse model, SITH-1 expression in the olfactory bulb led to apoptosis, decreased choline acetyltransferase expression in the brain, reduced hippocampal neurogenesis and increased depression-like behaviour ^92^. An earlier study showed similar findings, including that SITH-1-mediated olfactory bulb apoptosis led to HPA axis activation, and increased corticotropin-releasing hormone ^34^. Further, a study in humans showed that antibodies against HHV-6 were positively associated with higher cortisol ^93^. While we observed a weak positive trend between salivary HHV-6 DNA and a higher CAR AUC in the LC group, it did not reach statistical significance (r= 0.21, 95% CI = -0.11 to 0.49, p= 0.19, **Supplementary Figure 6E**). Taken together, these findings suggest that HHV-6 may play a direct, neuropathological role, though understanding the mechanistic relationship between HHV-6, stress, hormone responses and symptomology requires more work with a larger sample size.

While we did not observe associations between EBV and cortisol, higher waking salivary testosterone levels, particularly in males, were nominally associated with greater salivary EBV DNA. This aligns with the observation that males have higher anti-EBV IgA titres, consistent with more frequent mucosal reactivation, while females have higher anti-EBV IgG ^94^ titres, a pattern previously observed in a pre-print in people with LC ^30^.

This study has several limitations. The modest sample size limited the statistical power of some analyses and may increase the risk of selection bias. We also only captured a single period of hormonal and viral fluctuations in each participant. In addition, medication use was high, especially in individuals with LC. We accounted for this with strict post-hoc exclusions, but these relied largely on self-report data, supplemented with EMR review where possible. Although we centrifuged our samples to remove cells and calibrated our herpesvirus quantification to human nucleic acid quantity, it is possible that some herpesvirus DNA originated from latently infected cells. We further mitigated this with a stringent positivity threshold of LLOQ in ≥2 samples per individual.

Our study design used self-collected, unsupplemented saliva, which maximized convenience for participants and the breadth of compatible assays. Saliva was stored for a maximum of 120 hours in a home refrigerator, which may have led to some variability. Previous work has shown both DNA and cortisol are relatively stable for up to 5 days when stored at 4°C, ^95–97^ though testosterone and oestradiol may decrease more significantly after 3 days ^98^. Potential variation was further mitigated by testing multiple timepoints over two days and conducting analysis on aggregated values.

Additionally, the lack of a specialized storage buffer meant that reliable, comparative quantification of herpesvirus mRNA, which would have offered a conclusive method to confirm herpesvirus reactivation and further probe potentially relevant latency programs, was not possible. Indeed, it is possible that latent EBV might contribute to LC pathology, for example through reprogramming B-cells and driving autoimmunity ^99^, and HHV-6 pathology is often related to latency ^34,92^. Finally, our study did not examine VZV, HHV7 and HHV8, and we did not differentiate between HSV 1 and 2, nor HHV-6 A and B, which have distinct tropisms and potential pathobiology. Future longitudinal studies utilizing metagenomic and transcriptomic approaches are needed to more comprehensively assess the latency and reactivation stages of all HHVs, over the course of LC development, and their associations with LC pathology.

Our results suggest distinct roles for EBV and HHV-6 in LC. For EBV, the cumulative evidence from previous work suggests that reactivation before, during or shortly after acute SARS-CoV-2 infection may trigger pathological events that persist beyond acute infection ^19–21,23^. Until recently, it was widely considered that salivary shedding of HHV-6 is asymptomatic and not clinically important ^100^. While our findings recapitulate the known high prevalence of asymptomatic shedding of HHV-6, we additionally show an association between higher HHV-6 DNA copy number and symptom severity. This correlation suggests that persistent reactivation of HHV-6 in LC may actively contribute to pathology, which is supported by similar findings in ME/CFS ^44^. However, salivary HHV-6 has also been proposed as a biomarker of physiological or psychological stress ^101^. Thus, whether HHV-6 reactivation is a response to, or a trigger of, increased symptom severity remains to be investigated.

Off-label use of anti-herpes antivirals has been reported among those with LC ^102,103^, but the evidence base supporting their use is scarce, with only one clinical trial currently ongoing ^104^. Furthermore, no drugs have been approved to treat EBV or HHV-6 specifically in any setting, and there is limited-to-no evidence supporting the effectiveness of commonly prescribed nucleoside analogues such as valacyclovir ^105,106^. Development of effective antivirals, vaccines, and monoclonal antibodies for EBV and HHV-6 is needed to examine the role of these persistent herpesviruses in LC and other post-acute infection syndromes.

## Supporting information

STROBE checklist

## Data Availability

The qPCR analysis pipeline is available for public use and it is deposited, along with supporting files in GitHub (https://github.com/clairelaxton/Automated_qPCR_ETL_pipeline). The pre-specified rubric used for symptom survey encoding, statistical model results, and relevant de-identified data included in our analyses are included as supplementary files. Additional requests for data sharing should be directed to.

## Contributors

Conceptualization-CSL, AT, JS, JW, DP, AI; Methodology-CSL, AT, KW, SB, JS, JW, CM; Investigation-CSL, AT, LC, JS, SB, HN, ZL, GR, VF, CG, AL; Formal analysis-CSL, AT, KW; Visualization-CSL, AT, KW; Data Curation (including accessing and verifying underlying data-CSL, AT, LC, SB, JW, HN, ZL, WBH; Project administration-CSL, AT, LC, JS, JW, MD, BB, DP, AI; Supervision-LG, AI, DP; Resources-DP, AI; Funding acquisition-DP, AI; Writing - Original Draft-CSL, AT, KW; Writing - Review & Editing-all authors. All authors read and approved the final version of this manuscript.

## Acknowledgments

The authors would like to thank Isabel Ott for their help with study methodology and Courtney Fennell for their help refining our qPCR analysis pipeline. This work was supported in part by the Else Kröner Fresenius Prize for Medical Research 2023 (to A.I.), the Howard Hughes Medical Institute Collaborative COVID-19 Initiative (to A.I.), and the Howard Hughes Medical Institute (to A.I.), the Polybio Research Foundation (to A.I. and D.P.) and the Steven and Alexandra Cohen Foundation (to D.P.). National Institutes for Health (NIH) funding sources supported AT (NIAID F31AI181508), SB (NHLBI T32HL007974-24) and CM (NCATS TL1 TR001864).

