## Supplementary Figures and Tables for "Associations Between Salivary HHV-6 And Long COVID Severity in a Cross-Sectional, Case-Control Analysis"

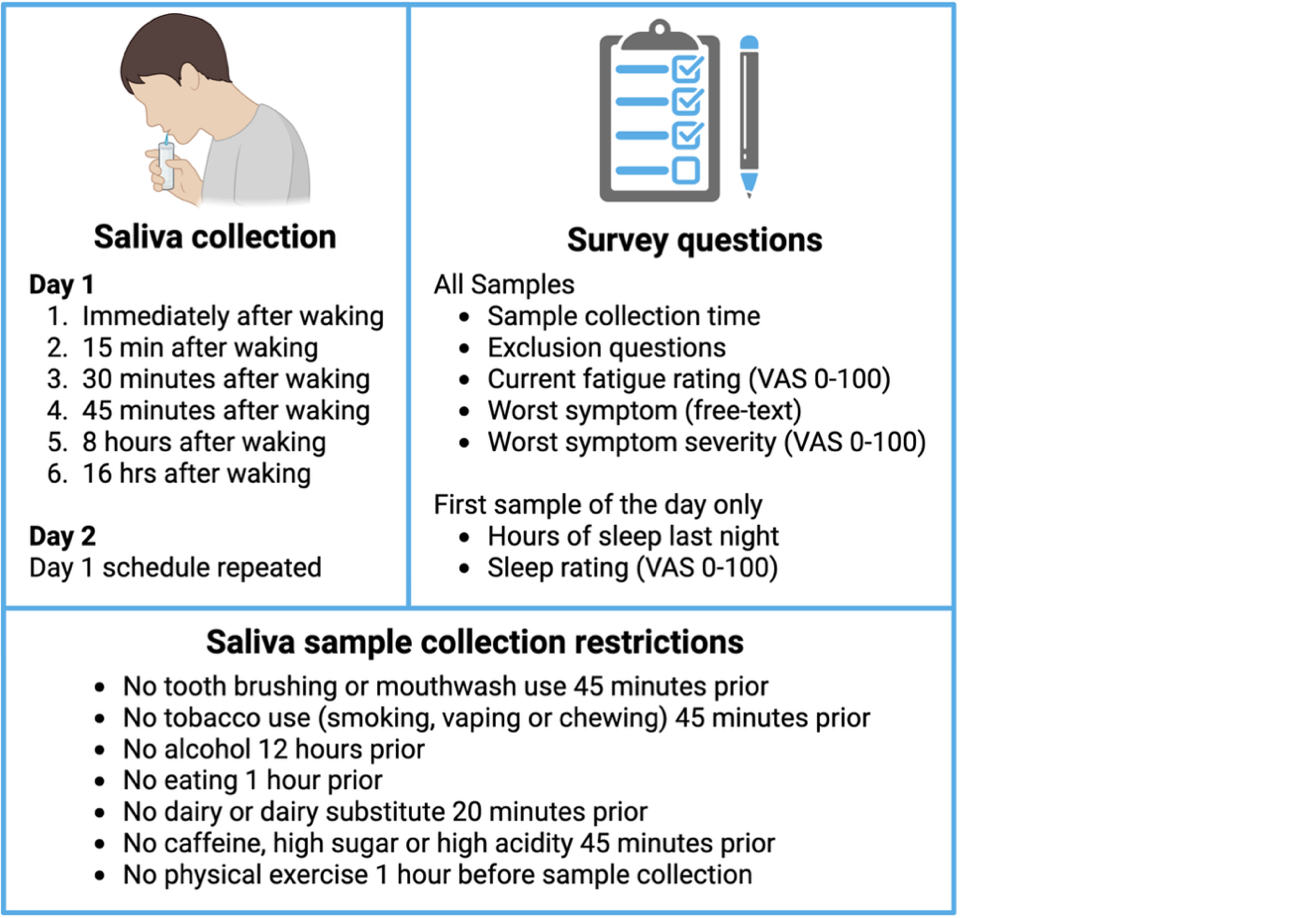

**Supplementary Figure 1: Saliva and daily sample survey collection schedule and restrictions.** As far as possible, surveys were collected at the same time as the saliva samples.

**Supplementary Table 1: Primers and probes used in this study.**

| **Target** | **Forward 5'-3' (final nM)** | **Reverse 5'-3' (final nM)** | **Probe 5'-3'**  **(final nM)** | **Source** |
| --- | --- | --- | --- | --- |
| **Herpesvirus multiplex** | | | | |
| **EBV** (BNRF1 gene) | GGAACCTGGTCATCCTTGC (300 nM) | ACGTGCATGGACCGGTTAAT (300 nM) | (FAM)-CGCAGGCACTCGTACTGCTCGCT-(IABKFQ) (200 nM) | ^107^ |
| **HCMV** (IE1 gene) | TGAAGCGCCGCATTGA (400 nM) | TGGCCCGTAGGTCATCCA (400 nM) | (HEX)-TCTGCATGAAGGTCTTTGCCCAGTACATCC-(IABKFQ) (200 nM) | Modified from ^108^ |
| **HSV** (1&2, (polymerase gene-coding region) | AGAGGGACATCCAGGACTTTG (200 nM) | GAGCTTGTAATACACCGTCAGG (200 nM) | (CY5)-CCGCCGAACTGAGCAGACACCCGCGC-(IABKFQ) (100 nM) | Modified from ^109^ |
| **HHV-6** (pan A/B U67-ORF gene) | CAAAGCCAAATTATCCAGAGCG (400 nM) | CGCTAGGTTGAGGATGATCGA (400 nM) | (ROX)-CACCAGACGTCACACCCGAAGGAAT-(IABRQSP) (200 nM) | ^110^ |
| **SARS-CoV-2 / RNAseP dual-plex** | | | | |
| **Human** (RNase P subunit 30 gene) | AGATTTGGACCTGCGAGCG (150 nM) | GAGCGGCTGTCTCCACAAGT (150 nM) | (CY5)-TTCTGACCTGAAGGCTCTGCGCG-(BHQ1) (200 nM) | ^111^ |
| **SARS-CoV-2** (N1 gene) | GACCCCAAAATCAGCGAAAT (500 nM) | TCTGGTTACTGCCAGTTGAATCTG (500 nM) | (FAM)-ACCCCGCATTACGTTTGGTGGACC-(BHQ1) (125 nM) | ^111^ |

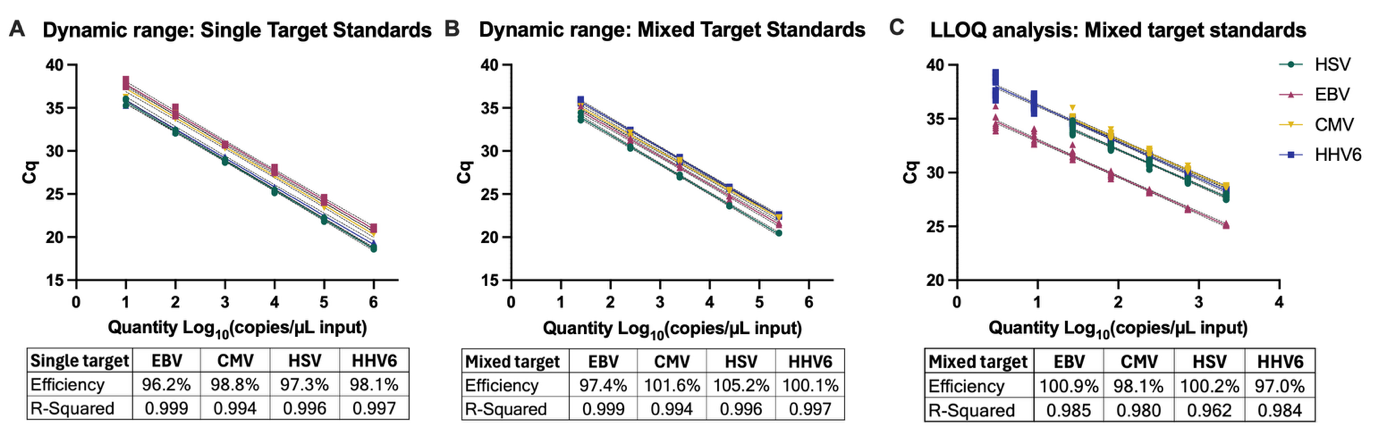

**Supplementary Figure 2: Representative standard curves for EBV, CMV, HSV and HHV-6 detection using quantitative genomic DNA standards.** Amplification was carried out with 1:10 serial dilutions of the quantitative gDNA for either one target of interest (**A**, single target, 200,000-2 copies/reaction), or a 1:1:1:1 mix of all targets (**B**, mixed target, 250,000-25 copies/reaction). Lower limits of quantification (LLOQ) were calculated (**C**, mixed-target, 2187-3 copies/reaction). LLOQ were 3 copies/µL input for EBV and HHV-6 and 27 copies/µL input for CMV and HSV. Efficiencies and R^2^ values from the simple linear regression (with 95% confidence interval, dashed lines) for each curve are shown in a panel below each experiment.

**Supplementary Table 2: Baseline demographic and clinical characteristics of age-sex-matched participants with Long COVID and Control participants.**

| **Demographics** | **Control** | **Long COVID** | **SMD** | **Δ** |
| --- | --- | --- | --- | --- |
| Age, median (IQR) | 36 ± 12 | 37 ± 13 | **0.366** | 0.53 |
| BMI, median (IQR) | 25 ± 6.4 | 23.5 ± 8.1 | -0.071 | -0.66 |
| % Female | 73.3% (33/45) | 73.3% (33/45) | 0 | 0 |
| Race: Asian | 13.3% (6/45) | 8.9% (4/45) | -0.142 | -4.4 |
| Race: Black or African American | 0% (0/45) | 2.2% (1/45) | **-** | NA |
| Race: Other | 6.7% (3/45) | 17.8% (8/45) | **0.344** | 11.1 |
| Race: White, other | 2.2% (1/45) | 2.2% (1/45) | 0 | 0 |
| Race: White, Hispanic | 0% (0/45) | 8.9% (4/45) | **-** | NA |
| Race: White, non-Hispanic | 77.8% (35/45) | 60% (27/45) | **-0.391** | -17.8 |
| **COVID-19 history (self-report)** | | | | |
| Previously had COVID-19 | 60% (27/45) | 100% (45/45) | **1.155** | 40 |
| Days since first COVID-19, median (IQR) | 877 ± 433.5 | 907 ± 776 | 0.186 | 77.27 |
| Days since last COVID-19, median (IQR) | 294 ± 263 | 270 ± 293 | **-0.364** | -77.25 |
| Positive test method: PCR | 44.4% (12/27) | 53.3% (24/45) | **-** | 8.9 |
| Positive test method: Antibody | 0% (0/27) | 8.9% (4/45) | **-** | NA |
| Positive test method: Antigen | 51.9% (14/27) | 17.8% (8/45) | **-** | -34.1 |
| Positive test method: None | 3.7% (1/27) | 20% (9/45) | **-** | 16.3 |
| Previously vaccinated against COVID-19 | 100% (45/45) | 100% (45/45) | **-** | 0 |
| Days since COVID vaccine, median (IQR) | 293 ± 531 | 280 ± 391 | -0.072 | -32.47 |
| **Reported medication use (last 3 months)** | | | | |
| Anti-hypertensives | 6.7% (3/45) | 4.4% (2/45) | -0.097 | -2.2 |
| Anti-platelet medication | 0% (0/45) | 8.9% (4/45) | **0.442** | 8.9 |
| Anti-psychotics | 0% (0/45) | 0% (0/45) | **-** | 0 |
| Antibiotics | 6.7% (3/45) | 2.2% (1/45) | **-0.217** | -4.4 |
| Antivirals | 4.4% (2/45) | 6.7% (3/45) | 0.097 | 2.2 |
| Blood thinners | 2.2% (1/45) | 13.3% (6/45) | **0.424** | 11.1 |
| Coenzyme Q10 or N-Acetyl L-Cysteine | 4.4% (2/45) | 51.1% (23/45) | **1.221** | 46.7 |
| Depression/anxiety treatment | 13.3% (6/45) | 40% (18/45) | **0.632** | 26.7 |
| Diabetes treatment | 2.2% (1/45) | 4.4% (2/45) | 0.124 | 2.2 |
| Steroids (inhaled/nasal) | 0% (0/45) | 0% (0/45) | **-** | 0 |
| NSAIDs | 33.3% (15/45) | 33.3% (15/45) | 0 | 0 |
| Nattokinase, serrapeptase or lumbrokinase | 2.2% (1/45) | 26.7% (12/45) | **0.742** | 24.4 |
| Opioids | 2.2% (1/45) | 0% (0/45) | **-0.213** | -2.2 |
| **Hormonal medications** | 22.2% (10/45) | 22.2% (10/45) | 0 | 0 |
| Hormonal intra-uterine device | 18.8% (3/16) | 31.2% (5/16) | **0.354** | 12.5 |
| Oral combined oestrogen-progestin | 31.2% (5/16) | 12.5% (2/16) | 0 | -18.8 |
| Oral progestin-only | 6.2% (1/16) | 0% (0/16) | **-0.577** | -6.2 |
| Other/unspecified | 6.2% (1/16) | 18.8% (3/16) | **0.577** | 12.5 |
| **Medical history (self-reported, past or current)** | | | | |
| Anxiety | 20% (9/45) | 55.6% (25/45) | **0.788** | 35.6 |
| Asthma | 11.1% (5/45) | 24.4% (11/45) | **0.354** | 13.3 |
| Auto-immune disease | 0% (0/45) | 0% (0/45) | - | - |
| Cancer | 4.4% (2/45) | 2.2% (1/45) | -0.124 | -2.2 |
| Chronic kidney disease | 0% (0/45) | 0% (0/45) | - | - |
| Chronic liver disease | 0% (0/45) | 0% (0/45) | - | - |
| Chronic obstructive pulmonary disease | 0% (0/45) | 0% (0/45) | - | - |
| Depression | 20% (9/45) | 35.6% (16/45) | **0.353** | 15.6 |
| Diabetes | 0% (0/45) | 0% (0/45) | - | - |
| Eating disorder | 6.7% (3/45) | 2.2% (1/45) | **-0.217** | -4.4 |
| Hormonal disorder | 2.2% (1/45) | 6.7% (3/45) | **0.217** | 4.4 |
| Hypertension | 4.4% (2/45) | 6.7% (3/45) | 0.097 | 2.2 |
| Obesity | 8.9% (4/45) | 6.7% (3/45) | -0.083 | -2.2 |
| Other chronic lung disease | 0% (0/45) | 4.4% (2/45) | **0.305** | 4.4 |
| Other neurological disease | 0% (0/45) | 2.2% (1/45) | **0.213** | 2.2 |
| Other psychiatric disease | 4.4% (2/45) | 4.4% (2/45) | 0 | 0 |
| Spinal cord injury | 0% (0/45) | 0% (0/45) | - | - |
| Stroke | 0% (0/45) | 0% (0/45) | - | - |
| Other comorbidities | 11.1% (5/45) | 35.6% (16/45) | **0.604** | 24.4 |
| *SMD= Standardized mean difference between groups (mean difference / pooled SD of the matched pair differences for continuous variables; difference in proportions / pooled binomial SD for categorical variables, reported per category). SMD ≥0.20 is indicative of meaningful imbalance and is shown in bold.*  *Δ= Absolute difference between groups (mean difference for continuous variables; percentage-point difference for categorical variables). NA= not available.* | | | | |

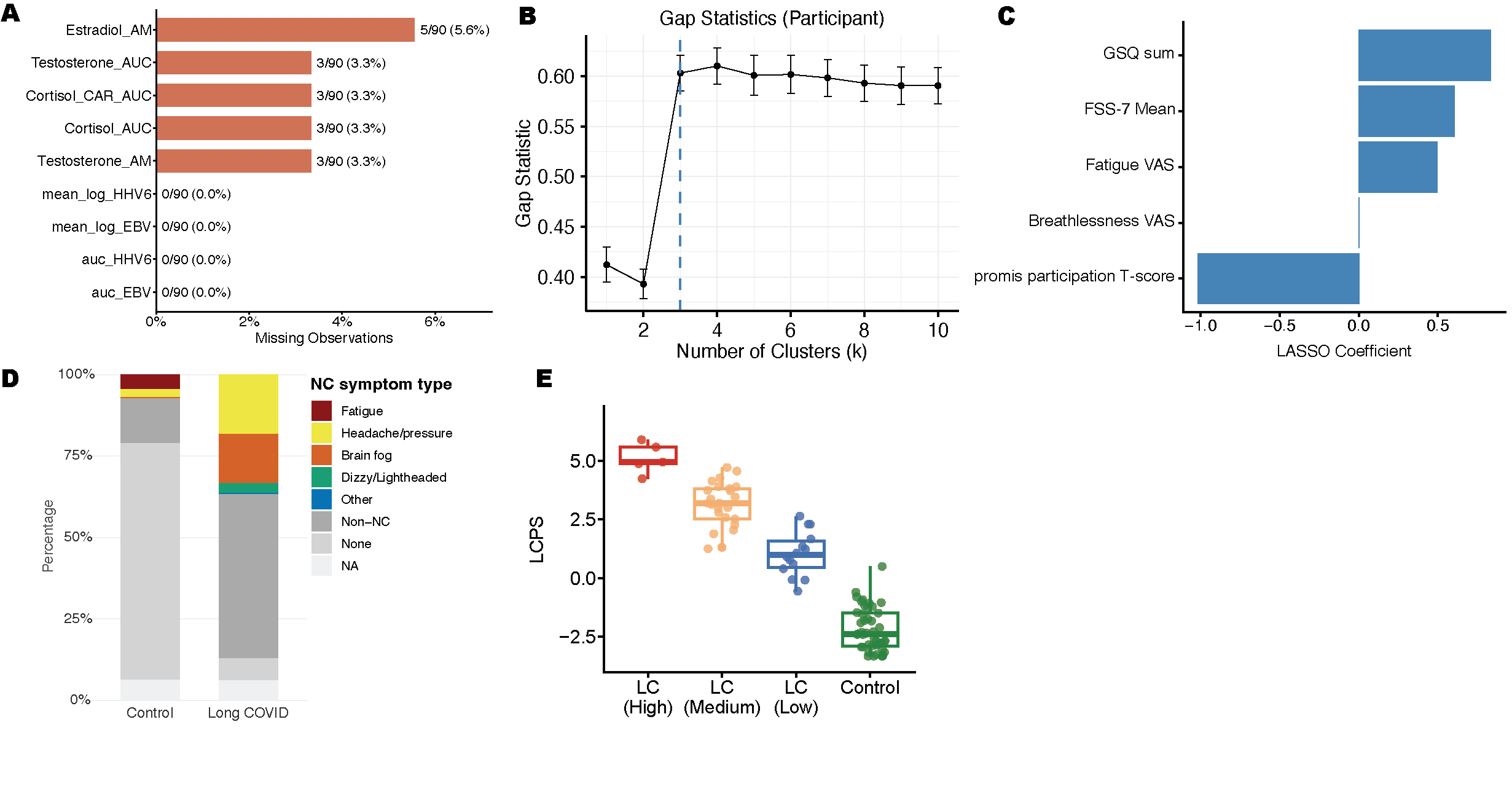

**Supplementary Figure 3: A.** Overall missingness of data for each participant. **B.** Gap statistics for determining the optimal number of clusters of participants for the GSQ-30 symptom clustering shown in **Figure 2A**. **C.** LASSO coefficients of the non-zero contributors to the LCPS model (GSQ Sum= Sum of General Symptom Questionnaire 30, FSS-7 Mean= mean fatigue severity scale score, VAS= visual analogue scale for fatigue and breathlessness, promis participation T-score= Adult PROMIS Ability to Participate in Social Roles and Activities short form. **D.** Incidence of neurocognitive (NC) symptom being reported by participants as the ‘worst symptom’ at a given timepoint, separated by subtypes (NA = survey question not answered). **E.** LCPS for each participant, plotted according to their GSQ-30 severity cluster**.**

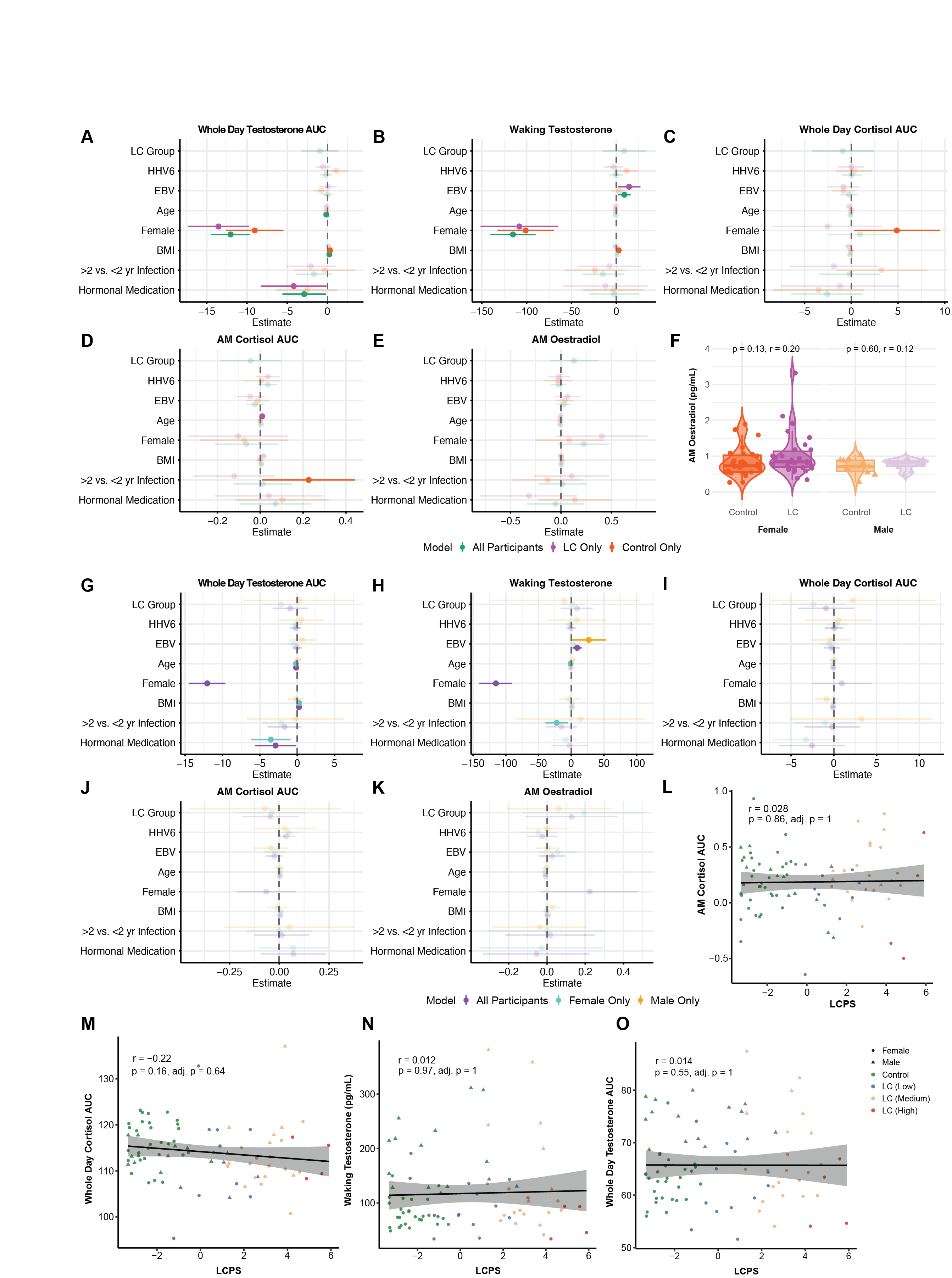

**Supplementary Figure 4: Relationships between salivary hormones, demographics, and Long COVID severity.** Forest plots showing the association between Whole Day Testosterone AUC (**A,G**), Waking Testosterone (**B,H**), Whole Day Cortisol AUC (**C,I**), Cortisol Awakening Response AUC (**D,J**) and Morning Oestradiol (**E,K**) and Long COVID status, EBV and HHV-6 mean log (viral copies) +1, adjusted for age, sex, hormonal medication, BMI, and time from first COVID infection, stratified by Long COVID status (**A-E**) and sex (**G-K**). Regression coefficients are reported with 95% confidence intervals, and those in bold are associations with nominal p<0.05. **F.** Comparison of single timepoint morning salivary Oestradiol levels by group, n=30 (LC Female), n=10 (LC Male), n=31 (Control Female), and n=11 (Control Male). Central lines indicate the group median, top and bottom lines indicate the 75^th^ and 25^th^ percentiles, respectively, the whiskers represent 1.5 x the interquartile range. Each dot represents one individual. Statistical significance of the difference in median values was determined using Wilcoxon ranked-sum test. **L-O**. Correlation analysis between Long COVID propensity score and Cortisol Awakening Response AUC (**L**), Whole Day Cortisol AUC (**M**), Waking Testosterone (**N**), or Whole Day Testosterone AUC (**O**). Spearman coefficients (R) are shown, with significance determined by linear regression adjusting age, sex and BMI. p <0.05 was considered significant. Holm’s procedure was used to correct for multiple comparisons. The black line depicts linear regression, and the shading shows the 95% CIs. Each dot represents one individual. n = 84 (41 LC, 43 control).

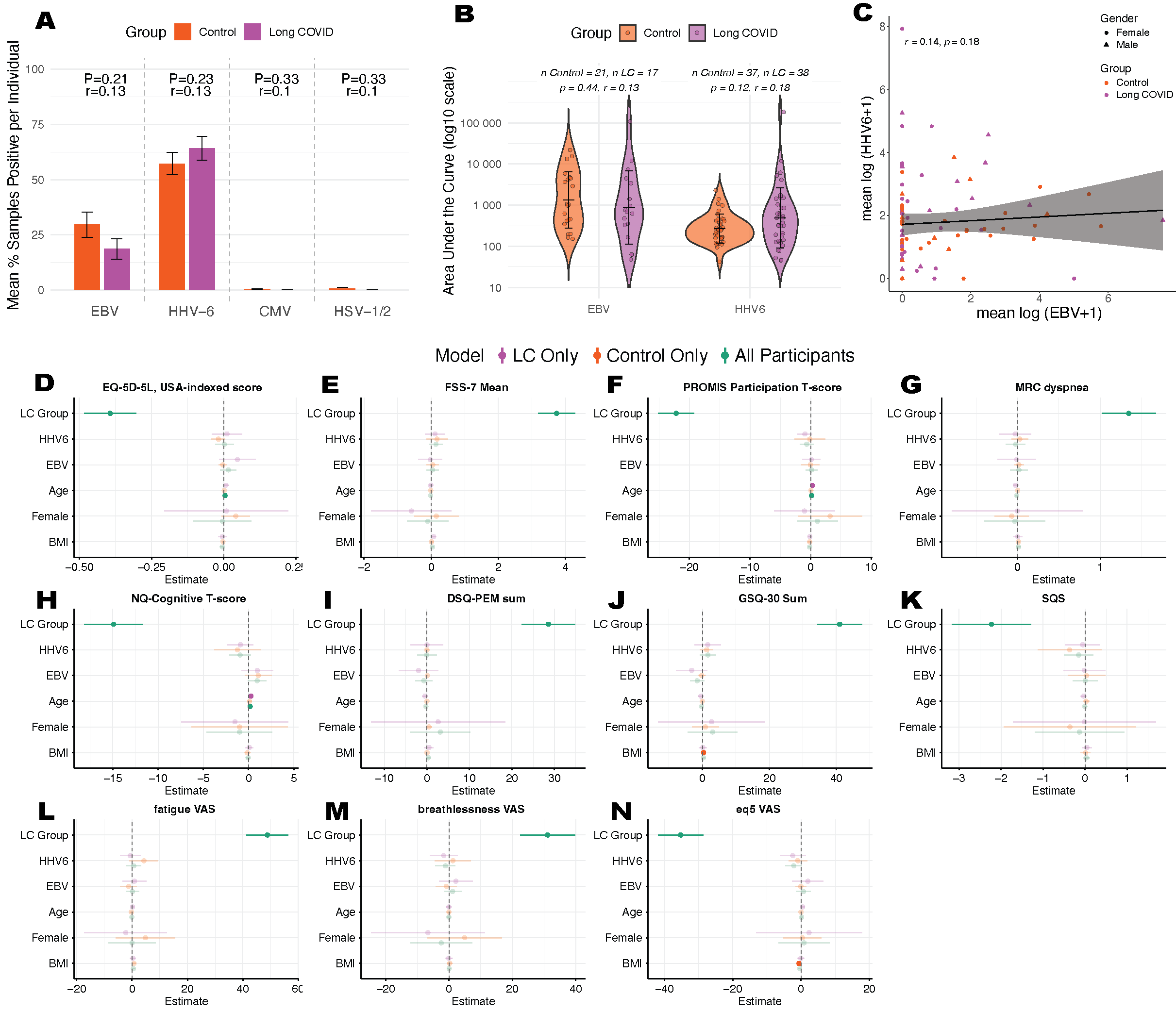

**Supplementary Figure 5: Relationships between detection of salivary herpesviruses, demographics, and Long COVID severity.** **A.** Proportion of samples per participant positive for each virus. Statistical differences tested using Fisher’s Exact test, OR= Odd’s Ratio, P<0.05 is significant. **B.** Comparison of area under the curve of viral DNA copies, averaged over 2 days, between participants with Long COVID and controls. Mean and SD are shown. Statistical differences tested using Wilcoxon rank-sum test, effect size reported as r (Z / √N), P<0.05 is significant. **C.** Spearman correlation between mean log (HHV-6 +1) and log (EBV+1) viral copies in saliva. P<0.05 is significant. **D-N.** Forest plots showing the association between health questionnaire outcomes and Long COVID (LC) status, EBV and HHV-6 mean log (viral copies) +1, adjusted for age, sex and BMI, and stratified by Long COVID status: quality of life (**D, N**), fatigue (**E, L**), ability to participate in daily activities (**F**), dyspnoea (**G, M**), perceived cognitive ability (**H**), post-exertional malaise (**I**), general somatic symptom burden (**J**) and sleep quality (**K**). Regression coefficients are reported with 95% confidence intervals and those in bold are associations with p<0.05, nominal significance did not survive Holm’s multiple correction.

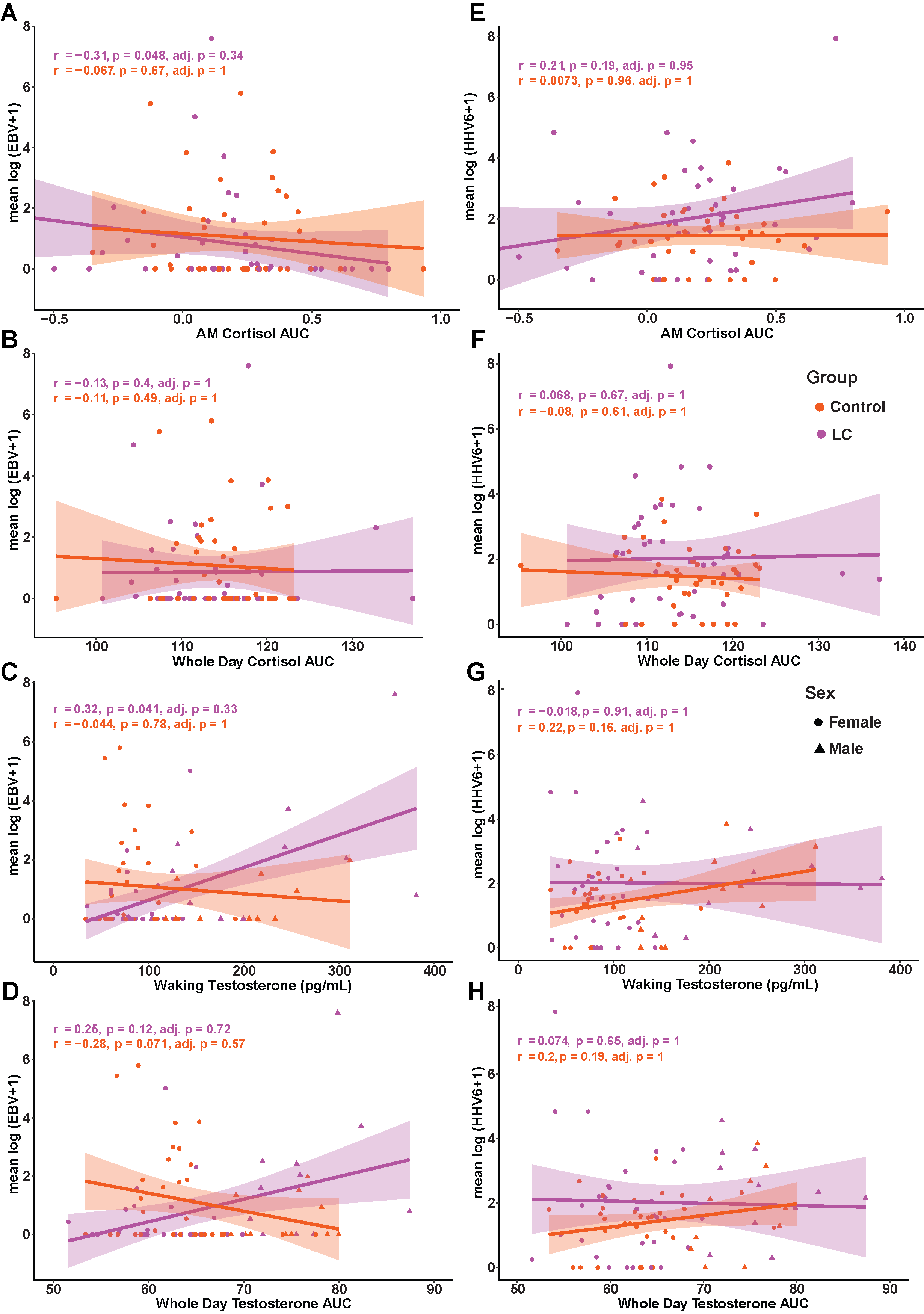

**Supplementary Figure 6: Relationships between salivary hormones and viral DNA detection. A-D.** Correlation analysis between salivary EBV DNA copies and Cortisol Awakening Response AUC (**A**), Whole Day Cortisol AUC (**B**), Waking Testosterone (**C**), and Whole Day Testosterone AUC (**D**) separated by group. **E-H.** Correlation analysis between salivary HHV-6 DNA copies and Cortisol Awakening Response AUC (**E**), Whole Day Cortisol AUC (**F**), Waking Testosterone (**G**), and Whole Day Testosterone AUC (**H**) separated by group. Spearman coefficients (R) are shown, with significance determined by linear regression. Holm's procedure was used to correct for multiple comparisons. p <0.05 was considered significant. The dark, solid lines depict linear regression, and the shading shows the 95% CIs. Each dot represents one individual. n=41 (LC) and n=43 (Control).
