## Supplementary Document 1 for "Associations Between Salivary HHV-6 And Long COVID Severity in a Cross-Sectional, Case-Control Analysis"

**Timepoint survey encoding protocol**

**Worst_symptom_category**

*Categorization will be completed by two independent encoders. Following this, any discrepancies between encoders, and all symptoms categorized as OTHER will be determined by a trained clinician.*

*Notes:*

- *If multiple symptoms listed, categorize only the first symptom listed (if first symptom is too difficult to interpret, move to second)*
- *If you are not sure what a participant meant, either because they used terms not listed in the scoring protocol, or their language was difficult to interpret, do not try to over-interpret or look up the term, categorize as OTHER.*
- MSK: musculoskeletal
  - *bone, muscle or joint pain, joint or muscle stiffness or weakness, joint swelling, feeling unsteady or weak when standing*
- PUL: pulmonary
  - *shortness of breath, breathlessness, difficulty breathing, cough, sputum (productive cough), upper respiratory symptoms including sore throat, congestion, runny nose*
- CV: cardiovascular
  - *abnormal heart rhythms (arrhythmia), palpitations, heart ‘skipping a beat’, chest pain*
- NC: neurocognitive
  - *poor cognition and focus e.g. brain fog, disorientation, memory problems, tiredness, pain if with reference to tingling, numbness, burning, "pins and needles”, tension headaches, migraines, tremors, feeling lightheaded, drowsy or groggy, fainting, headaches, mentions of orthostatic hypotension or POTS, loss of taste or smell*
- PSYCH: Psychiatric
  - *symptoms relating to mood, sleep, depression/anxiety or other psychiatric symptoms, insomnia*
- SH: skin and hair
  - *lesions, rashes, lumps, tightness, itching, dermatitis or heightened skin allergies, unexplained hair loss*
- EE: Eyes and ears
  - *unpleasant sensations in EE, vision changes e.g. seeing spots or having tunnel vision, increased shortsightedness or blurred vision, ear fullness, blocked ears, tinnitus, loss of hearing****,*** *hypersensitivity to visual/audible stimuli*
- GI: gastrointestinal
  - *vomiting, diarrhea, nausea), bloating, burping, changes in appetite indigestion, acid reflux, abdominal pain*
- UG: Urogenital
  - *symptoms relating to pain, difficulty or alterations in urination or menstruation, vaginitis, discharge, sexual dysfunction*
- WB: Whole body
  - *post exertional malaise or PEM, whole body fatigue or pain, fever, chills, sweating, weight loss (or gain)*
- EXT: Extremities
  - *swelling of legs or hands; swollen, raised or painful veins*
- OTHER: If category not available or interpretation is difficult
  - *cannot be easily categorized*
- None: Reports “no symptoms”, “none” or related.
  - *Or if reported symptom is clearly unrelated to general health (e.g. pain from a specified injury, symptoms specifically due to lack of food/drink during sample collection), also mark to exclude symptom score in such case.*

**Secondworst_symptom_catagory**

*If multiple symptoms listed from multiple categories, categorize the second symptom listed.*

**Pain**

- Yes
- No

*If yes:*

**Pain_area**

- Head and neck
- Chest
- Abdomen
- MSK *(bones, muscles, joints)*
- Nerves *(if accompanied by tingling/burning or temperature/light touch sensitivity)*
- Other
- Whole_body

**Exclude_symptom_score**

*If symptoms are too ambiguous to understand, clearly unrelated to general health (e.g. pain from a specified injury, symptoms specifically due to lack of food/drink during sample collection), or if a score has been given but no symptom is noted, or a symptom given but not score noted.*

- Yes
- Leave blank if no
